# Identifying outcomes for evaluating the impact of pharmacist prescribing: A rapid overview of reviews

**DOI:** 10.1101/2025.09.04.25334869

**Authors:** Ahmed Hassan Ali, Anais Essilini, Aaron Daunt, Michelle Flood, Caroline McCarthy, Judith Strawbridge, Barbara Clyne, Frank Moriarty

**Affiliations:** School of Pharmacy and Biomolecular Sciences, Royal College of Surgeons in Ireland (RCSI) University of Medicine and Health Sciences; RCSI PPI Ignite Network, Office of Research and Innovation, RCSI University of Medicine and Health Sciences; Department of General Practice, RCSI University of Medicine and Health Sciences; Department of Public Health and Epidemiology, RCSI University of Medicine and Health Sciences

**Keywords:** Pharmacist, Role Expansion, Prescribing, Common Conditions, Minor Ailments

## Abstract

**Objectives:** Given the expansion of pharmacist prescribing, this study aims to identify and categorise outcomes reported in research evaluating impacts of pharmacist prescribing specifically for minor ailments (or common conditions) and in other broader contexts.

**Methods:** A rapid overview of reviews was conducted, searching PubMed, Embase, Cochrane Database of Systematic Reviews, and Epistemonikos using keywords relating to pharmacy/pharmacists, prescribing or minor ailments schemes, and evidence syntheses. Reviews evaluating the impact of any aspect of pharmacist prescribing were included if they reported ≥1 outcome assessing the impact of pharmacist prescribing. Data extraction focused on the clinical settings, disease areas/conditions, prescribing models and reported outcomes.

**Results:** Of 43 reviews included, 14 reported on outcomes of pharmacist prescribing in minor ailments, and 35 reviews in broader contexts (some reporting both). Outcomes were categorised as clinical, drug-related/prescribing, patient-reported/experience, and economic/other outcomes. For minor ailments, 14 outcomes were identified, most frequently clinical cure or symptom resolution, and cost of service delivery (both reported in n=8 reviews). In other prescribing contexts, 12 outcomes were reported, with general satisfaction being the most common (n=18), followed by clinical effectiveness and healthcare resource use (both n=15).

**Conclusions:** Capturing impact of pharmacist prescribing requires assessment of outcomes across multiple dimensions.

## Background

Global healthcare systems are experiencing a shortage of skilled health care professionals, which is burdening primary care services, causing delays in access and increasing pressure on emergency departments and secondary care.^1, 2, 3^ Therefore various countries, including the United Kingdom (UK), Canada, the United States of America (USA), Australia, New Zealand, and France, have implemented strategies to expand the roles of healthcare professionals through task-shifting approaches.^1, 2^ These measures include granting prescribing authority to additional healthcare professionals, such as pharmacists to improve patient care outcomes without compromising safety, improve patient access to treatment, and make better use of existing clinical expertise.^3, 4^

Pharmacist prescribing varies significantly between and even within countries, particularly with respect to terminology and the extent of prescriptive authority.^5, 6^ In the UK, independent pharmacist prescribers are legally authorised to assess patients, make clinical decisions, and prescribe autonomously within their clinical competence without direct supervision.^7^ In Canada and the USA, prescribing authority differs by province and state, ranging from no prescribing rights to initiating prescriptions for specific indications and independent authority.^5, 6^ In other countries like Brazil and New Zealand, dependent or collaborative models apply following a prior medical diagnosis and collaborative agreements with medical prescribers.^6, 8^ In Ireland, a phased implementation of pharmacist prescribing is proposed, starting with common conditions in community pharmacy and progressing to full prescribing authority across primary and secondary care, in line with pharmacists’ scope of practice and competence.^9^

Expanding pharmacists’ roles through prescriptive authority is intended to alleviate the burden on overextended healthcare systems and enhance access to care.^1, 2^ It has also been linked with a range of positive clinical impacts, such as better glycaemic and hypertension management, improved quality of life for patients with chronic conditions, and enhanced prescribing practices (e.g. medicines optimisation/deprescribing).^2, 10, 11, 12^ However, evaluation of such role expansion to assess the effects (both intended and unintended) is important to assess whether changes are beneficial. Understanding the outcomes that have been used to evaluate the impact of pharmacist prescribing in a structured way can add coherence to the literature to-date and support alignment of future research on this topic. Therefore, this review aims to identify and categorise the outcomes synthesised in the literature on evaluation of the impact of pharmacist prescribing, guiding the identification of core outcomes and providing a foundation for more robust service evaluation.

## Methods

This was a rapid overview of reviews, for which a protocol was registered on Open Science Framework at the stage of title/abstract screening,^13^ and reported in line with the preferred reporting items for overviews of reviews (PRIOR) statement.^14^ A rapid overview of reviews was selected because of the need to identify outcomes within a constrained time period within a broader research project, and because synthesised evidence on evaluations of pharmacist prescribing was appropriate for identifying outcomes used.^15^

### Eligibility criteria and search strategy

Table 1 outlines the full eligibility criteria. A search string was developed reflecting the eligibility criteria, combining keywords and subject headings relating to the concepts of pharmacy or pharmacists, prescribing or minor ailments schemes, and systematic or other reviews. The search was conducted from database inception to 29^th^ November 2024 in the following databases: PubMed, Embase, Cochrane Database of Systematic Reviews, and Epistemonikos. Full search strategies are included in Appendix 1.

**Table 1.** Inclusion and exclusion criteria.

| Criteria | Inclusion | Exclusion |
| --- | --- | --- |
| Prescribing definition/context | Involved initiation, dose adjustment, substitution, extension, continuation, or discontinuation/deprescribing that is implemented by the pharmacist either independently or under a formal agreement with other independent prescribers.<br>Included substitution of one medicine with another containing a different active pharmaceutical ingredient (which may be in the same drug class or from a different one e.g. substituting a H2 receptor antagonist for a proton pump inhibitor).<br>Included prescribing in the context of common conditions, such as a minor ailments or common conditions schemes i.e. those where a pharmacist can supply a medicine that is normally prescription only to a patient who presents with a specified condition, usually of a minor or self-limiting nature.<br>Included other scenarios where a pharmacist was supplying prescription only medicines without a prescription from a prescriber.<br>Reviews focusing on prescribing by non-medical healthcare professionals in general but included studies focusing specifically on pharmacist prescribing if it was possible to identify pharmacist-specific evidence within these studies. | Reviews focusing on pharmacist supply where a medicine which is specifically referred to as non-prescription, and vaccine supply and administration. |
| **Outcomes of interests** | Must consider at least one outcome reflecting the impact or effect of pharmacist prescribing. | Reviews focusing only on other outcomes that did not reflect the impact or effect of pharmacist prescribing e.g. attitudes towards pharmacist prescribing. |
| **Reviews type and methodology** | • Systematic reviews, scoping reviews, or overviews of reviews, focusing on evaluating the effect/impact of expanded pharmacist roles involving any aspect of prescribing.<br>• Reviews that only included some studies relating to the aim were included and only details of relevant studies were extracted.<br>• Reviews were required to include a clear description of their search strategy, ideally including full search strings and database details (at least 2 databases), a structured approach to screening and reviewing papers for inclusion involving at least two reviewers, and a structured synthesis of the results which may or may not involve quantitative synthesis. |  |

### Screening and full-text review

Records from searches were uploaded to Covidence (Veritas Health Innovation, Melbourne, Australia), where duplicate records were removed. Screening was then conducted to exclude records clearly not meeting eligibility criteria based on title and abstract. Remaining records progressed to full-text review, where they were assessed against the eligibility criteria. For both stages, each record was reviewed independently by two reviewers (of AE, AD, AHA, or FM), with disagreements resolved by consensus.

### Data extraction and synthesis

For included reviews, data extraction focused on the review characteristics e.g. study details, type of pharmacist role examined (for common condition/minor ailments and other pharmacist prescribing), any restriction in setting, specific patient population(s), or other aspects, extracting details of the outcomes used to assess effect/impact. A narrative synthesis was conducted, mapping outcomes to the taxonomy developed by Dodd *et al*. ^16^ For the purpose of the current review, which aimed to explore and describe the body of evidence concerning the relevant outcomes of pharmacist prescribing, results of all included reviews were considered regardless of any overlap, and no quality appraisal was conducted to enable rapid identification of outcomes used.^17, 18, 19^

## Results

Of 1,727 articles initially retrieved, 1,152 articles underwent title/abstract screening where, 1,056 were excluded for not meeting the inclusion criteria. Ninety-six full texts were reviewed and 43 reviews fulfilled the eligibility criteria (see Figure 1).

**Figure 1.**
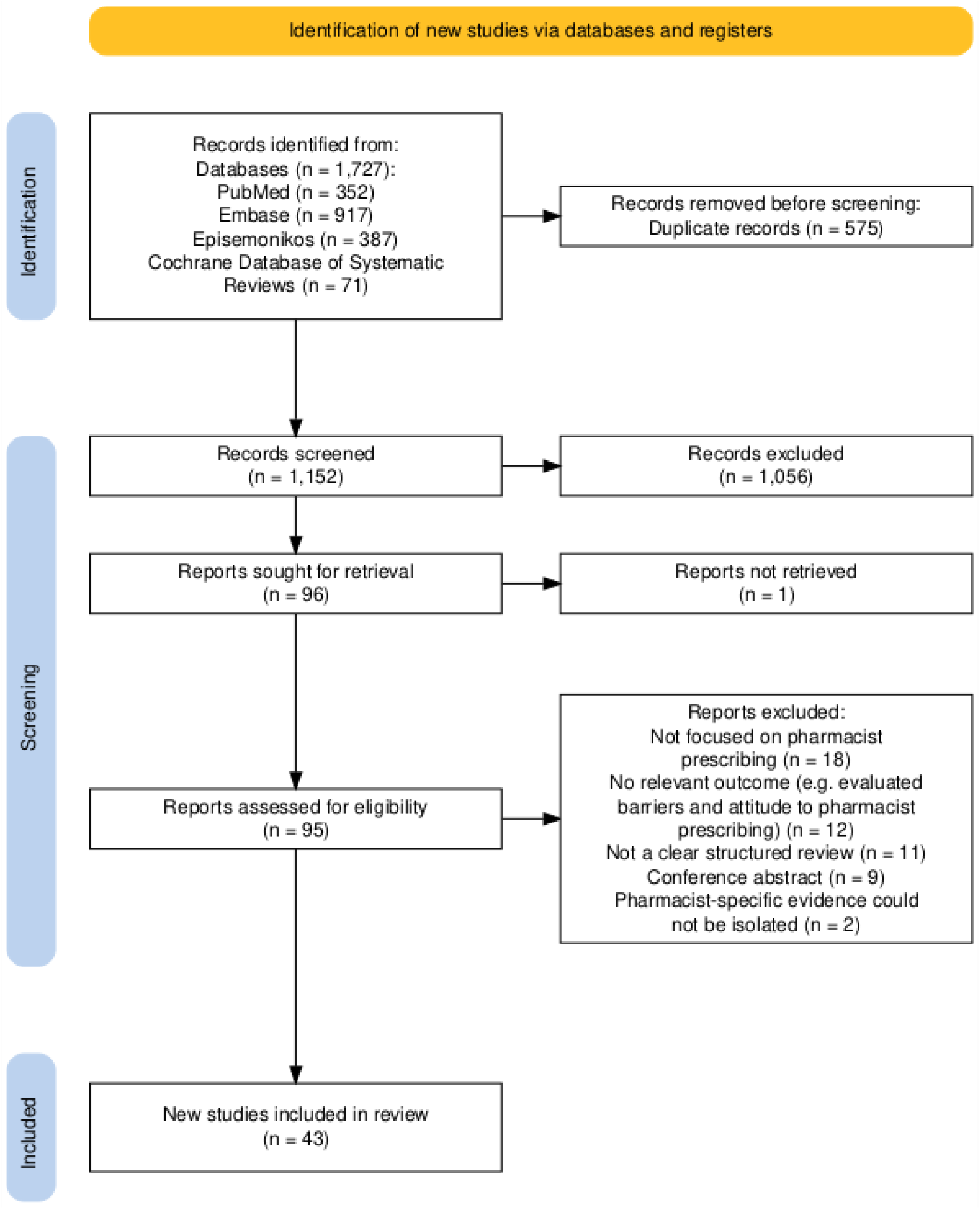
Flow chart of process of including eligible reviews within the overview of reviews

### Characteristics of the studies

The included reviews, published between 2008 and 2024, primarily consisted of systematic (n=30) and scoping reviews (n=10), with 3 overviews of reviews. The reviews included studies conducted across various healthcare settings, including community pharmacies, primary and secondary care, emergency departments, outpatient clinics, and nursing homes (Appendix 2). The types of prescribing examined across the included reviews encompassed varying levels of pharmacist authority in medication management. These included dependent, collaborative, supplementary, and independent prescribing and non-medical switching. The included studies covered diverse disease areas such as cardiovascular disease, diabetes, mental health, chronic kidney disease, oncology, perioperative care, and self-limiting and minor ailments.

Overall, 14 out of 43 reviews reported outcomes assessing the impact of pharmacist prescribing for minor ailments. Thirty-five reviews examined the impact of pharmacist prescribing for management of acute and chronic diseases beyond minor ailments (six covered prescribing in both minor ailments and other prescribing contexts).

### Outcomes for prescribing for minor ailments and other contexts

Tables 2 and 3 summarise the outcomes that were identified as assessing the impact of pharmacist prescribing for management of minor ailments and other pharmacist prescribing (with review-level results in Appendices 3-6). Outcomes were grouped into four categories: clinical; drug-related and prescribing; patient reported and experience; and economic and other related outcomes.

**Table 2.**
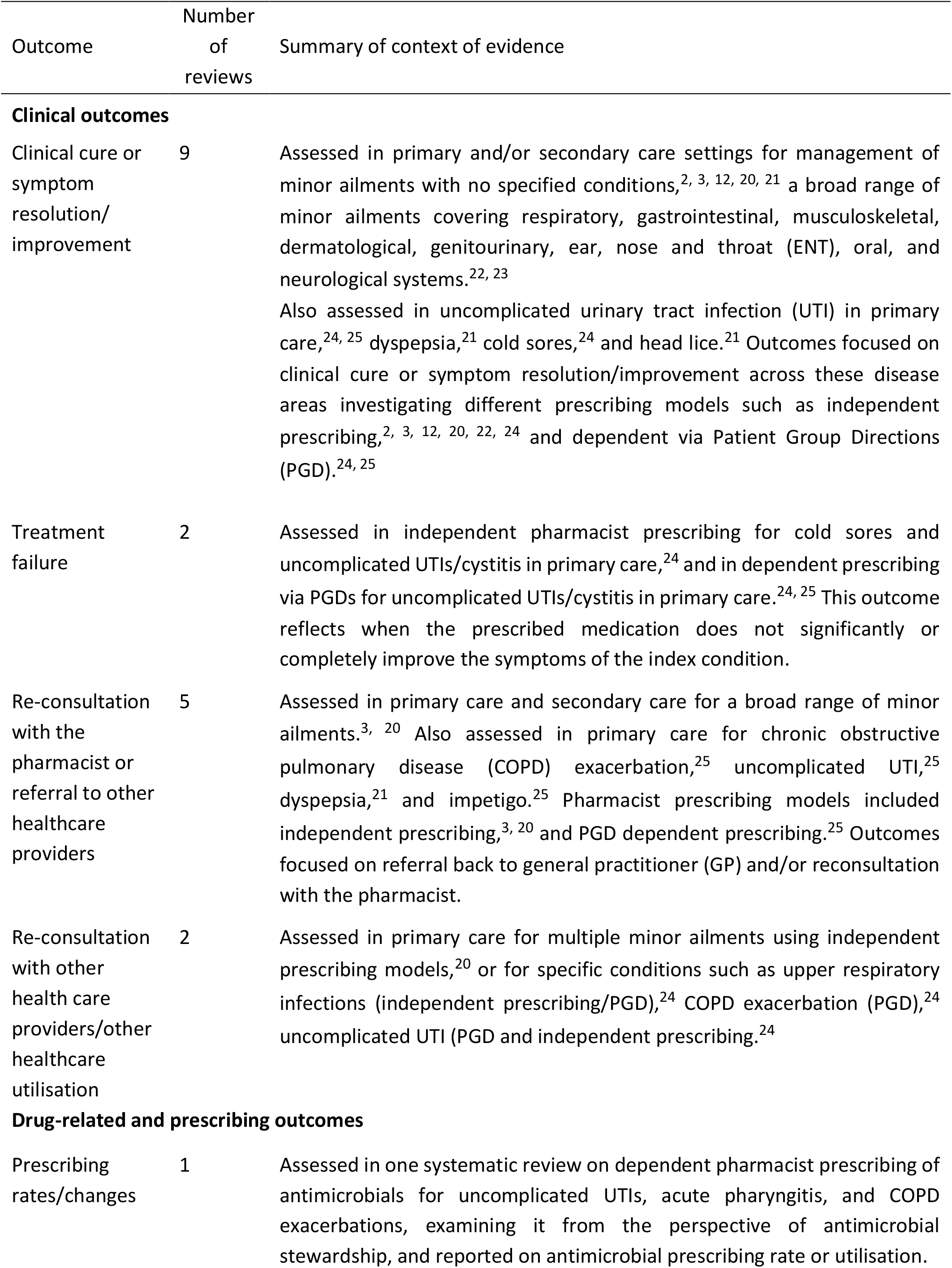
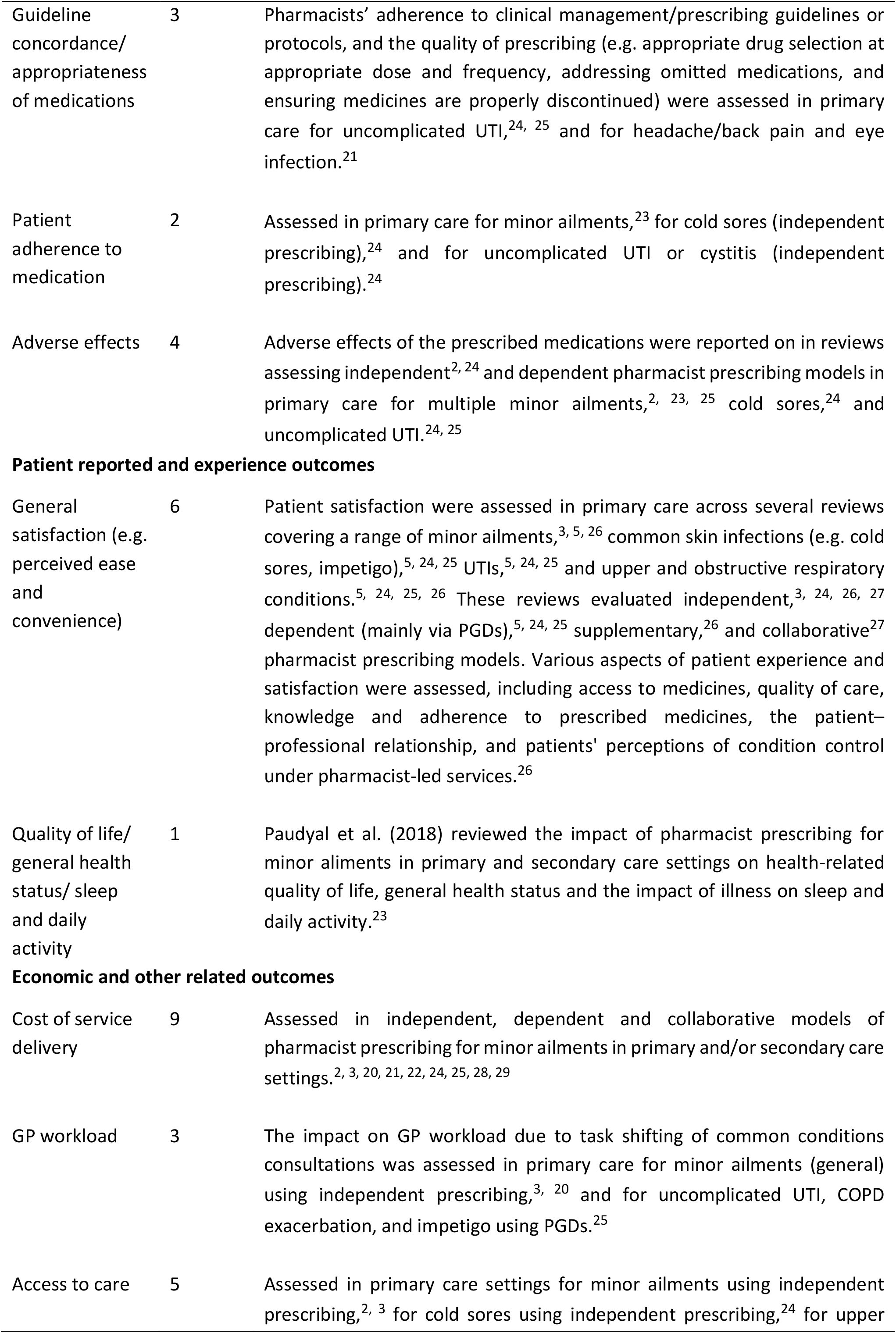
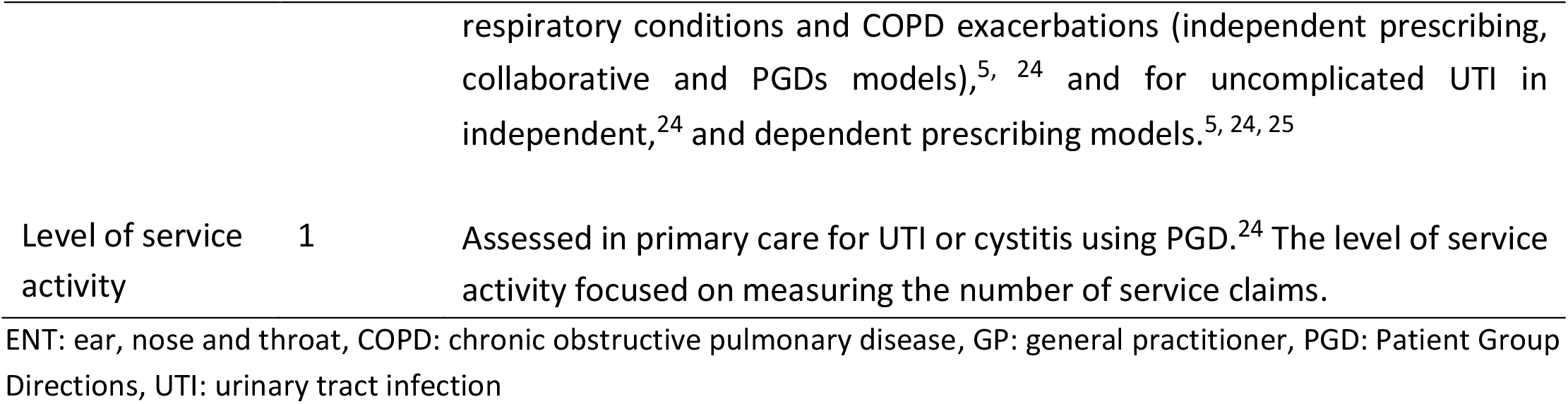
Outcomes reported for evaluating pharmacist prescribing for minor ailments (n=14 reviews)

**Table 3.**
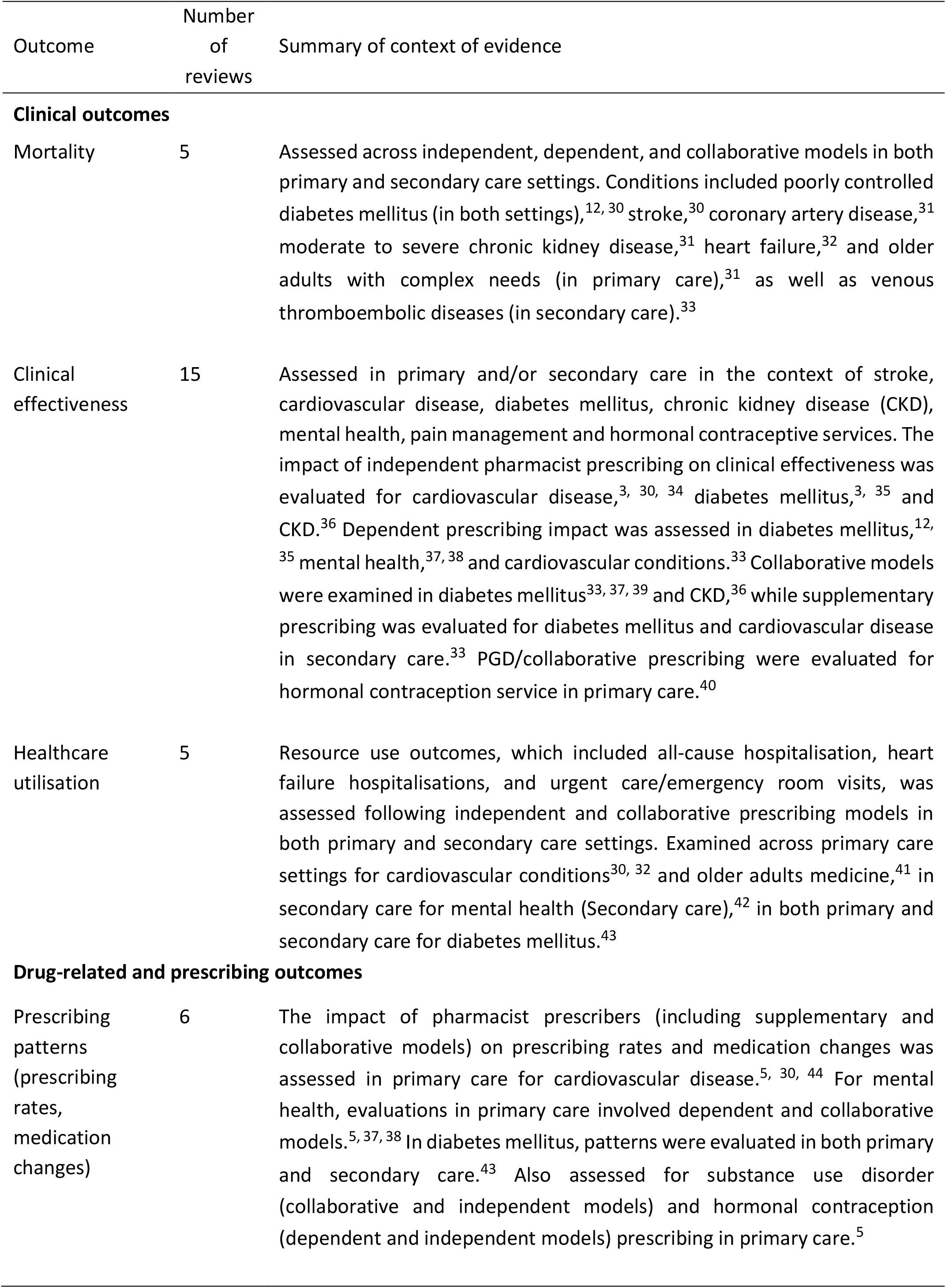
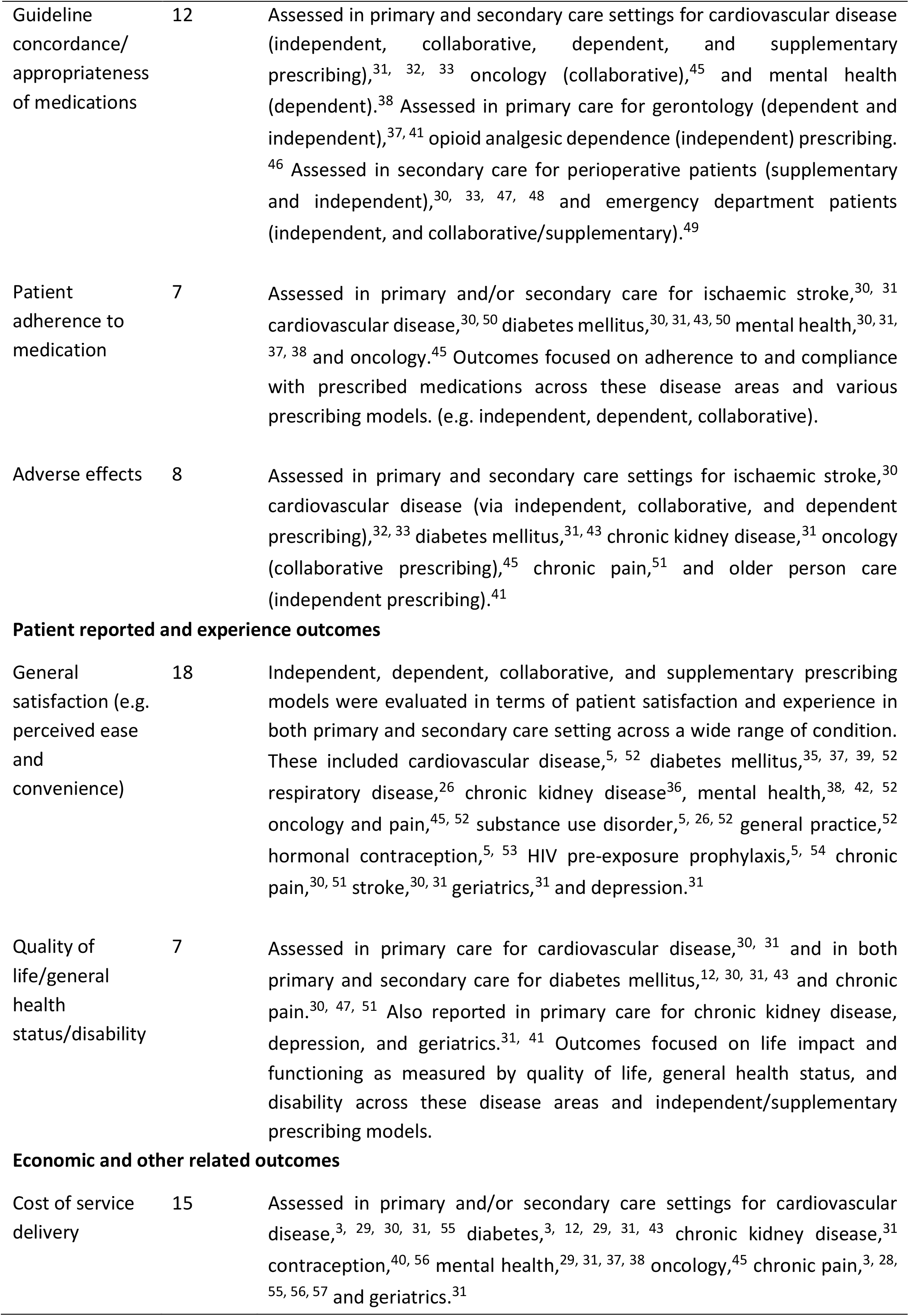
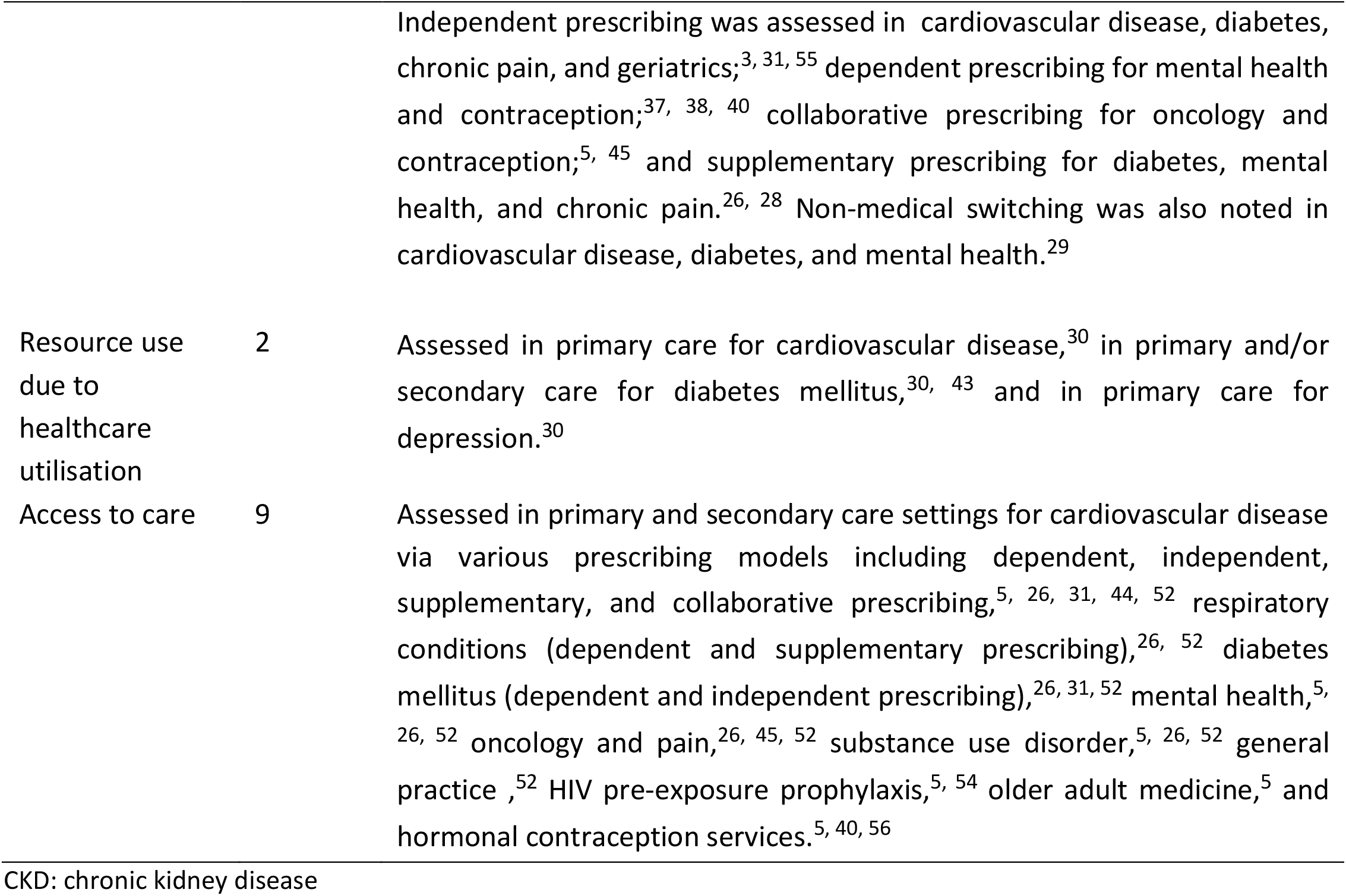
Outcomes for evaluating Other Pharmacist Prescribing (n=35 reviews)

| Outcome | Number of reviews | Summary of context of evidence |
| --- | --- | --- |
| **Clinical outcomes** |  |  |
| Mortality | 5 | Assessed across independent, dependent, and collaborative models in both primary and secondary care settings. Conditions included poorly controlled diabetes mellitus (in both settings),<sup>12, 30</sup> stroke,<sup>30</sup> coronary artery disease,<sup>31</sup> moderate to severe chronic kidney disease,<sup>31</sup> heart failure,<sup>32</sup> and older adults with complex needs (in primary care),<sup>31</sup> as well as venous thromboembolic diseases (in secondary care).<sup>33</sup> |
| Clinical effectiveness | 15 | Assessed in primary and/or secondary care in the context of stroke, cardiovascular disease, diabetes mellitus, chronic kidney disease (CKD), mental health, pain management and hormonal contraceptive services. The impact of independent pharmacist prescribing on clinical effectiveness was evaluated for cardiovascular disease,<sup>3, 30, 34</sup> diabetes mellitus,<sup>3, 35</sup> and CKD.<sup>36</sup> Dependent prescribing impact was assessed in diabetes mellitus,<sup>12, 35</sup> mental health,<sup>37, 38</sup> and cardiovascular conditions.<sup>33</sup> Collaborative models were examined in diabetes mellitus<sup>33, 37, 39</sup> and CKD,<sup>36</sup> while supplementary prescribing was evaluated for diabetes mellitus and cardiovascular disease in secondary care.<sup>33</sup> PGD/collaborative prescribing were evaluated for hormonal contraception service in primary care.<sup>40</sup> |
| Healthcare utilisation | 5 | Resource use outcomes, which included all-cause hospitalisation, heart failure hospitalisations, and urgent care/emergency room visits, was assessed following independent and collaborative prescribing models in both primary and secondary care settings. Examined across primary care settings for cardiovascular conditions<sup>30, 32</sup> and older adults medicine,<sup>41</sup> in secondary care for mental health (Secondary care),<sup>42</sup> in both primary and secondary care for diabetes mellitus.<sup>43</sup> |
| **Drug-related and prescribing outcomes** |  |  |
| Prescribing patterns (prescribing rates, medication changes) | 6 | The impact of pharmacist prescribers (including supplementary and collaborative models) on prescribing rates and medication changes was assessed in primary care for cardiovascular disease.<sup>5, 30, 44</sup> For mental health, evaluations in primary care involved dependent and collaborative models.<sup>5, 37, 38</sup> In diabetes mellitus, patterns were evaluated in both primary and secondary care.<sup>43</sup> Also assessed for substance use disorder (collaborative and independent models) and hormonal contraception (dependent and independent models) prescribing in primary care.<sup>5</sup> |
| Guideline concordance/<br>appropriateness<br>of medications | 12 | Assessed in primary and secondary care settings for cardiovascular disease (independent, collaborative, dependent, and supplementary prescribing),<sup>31, 32, 33</sup> oncology (collaborative),<sup>45</sup> and mental health (dependent).<sup>38</sup> Assessed in primary care for gerontology (dependent and independent),<sup>37, 41</sup> opioid analgesic dependence (independent) prescribing.<sup>46</sup> Assessed in secondary care for perioperative patients (supplementary and independent),<sup>30, 33, 47, 48</sup> and emergency department patients (independent, and collaborative/supplementary).<sup>49</sup> |
| Patient adherence to medication | 7 | Assessed in primary and/or secondary care for ischaemic stroke,<sup>30, 31</sup> cardiovascular disease,<sup>30, 50</sup> diabetes mellitus,<sup>30, 31, 43, 50</sup> mental health,<sup>30, 31, 37, 38</sup> and oncology.<sup>45</sup> Outcomes focused on adherence to and compliance with prescribed medications across these disease areas and various prescribing models. (e.g. independent, dependent, collaborative). |
| Adverse effects | 8 | Assessed in primary and secondary care settings for ischaemic stroke,<sup>30</sup> cardiovascular disease (via independent, collaborative, and dependent prescribing),<sup>32, 33</sup> diabetes mellitus,<sup>31, 43</sup> chronic kidney disease,<sup>31</sup> oncology (collaborative prescribing),<sup>45</sup> chronic pain,<sup>51</sup> and older person care (independent prescribing).<sup>41</sup> |
| Patient reported and experience outcomes |  |  |
| General satisfaction (e.g. perceived ease and convenience) | 18 | Independent, dependent, collaborative, and supplementary prescribing models were evaluated in terms of patient satisfaction and experience in both primary and secondary care setting across a wide range of condition. These included cardiovascular disease,<sup>5, 52</sup> diabetes mellitus,<sup>35, 37, 39, 52</sup> respiratory disease,<sup>26</sup> chronic kidney disease<sup>36</sup>, mental health,<sup>38, 42, 52</sup> oncology and pain,<sup>45, 52</sup> substance use disorder,<sup>5, 26, 52</sup> general practice,<sup>52</sup> hormonal contraception,<sup>5, 53</sup> HIV pre-exposure prophylaxis,<sup>5, 54</sup> chronic pain,<sup>30, 51</sup> stroke,<sup>30, 31</sup> geriatrics,<sup>31</sup> and depression.<sup>31</sup> |
| Quality of life/general health status/disability | 7 | Assessed in primary care for cardiovascular disease,<sup>30, 31</sup> and in both primary and secondary care for diabetes mellitus,<sup>12, 30, 31, 43</sup> and chronic pain.<sup>30, 47, 51</sup> Also reported in primary care for chronic kidney disease, depression, and geriatrics.<sup>31, 41</sup> Outcomes focused on life impact and functioning as measured by quality of life, general health status, and disability across these disease areas and independent/supplementary prescribing models. |
| Economic and other related outcomes |  |  |
| Cost of service delivery | 15 | Assessed in primary and/or secondary care settings for cardiovascular disease,<sup>3, 29, 30, 31, 55</sup> diabetes,<sup>3, 12, 29, 31, 43</sup> chronic kidney disease,<sup>31</sup> contraception,<sup>40, 56</sup> mental health,<sup>29, 31, 37, 38</sup> oncology,<sup>45</sup> chronic pain,<sup>3, 28, 55, 56, 57</sup> and geriatrics.<sup>31</sup> |
|  |  | Independent prescribing was assessed in cardiovascular disease, diabetes, chronic pain, and geriatrics;<sup>3, 31, 55</sup> dependent prescribing for mental health and contraception;<sup>37, 38, 40</sup> collaborative prescribing for oncology and contraception;<sup>5, 45</sup> and supplementary prescribing for diabetes, mental health, and chronic pain.<sup>26, 28</sup> Non-medical switching was also noted in cardiovascular disease, diabetes, and mental health.<sup>29</sup> |
| Resource use due to healthcare utilisation | 2 | Assessed in primary care for cardiovascular disease,<sup>30</sup> in primary and/or secondary care for diabetes mellitus,<sup>30, 43</sup> and in primary care for depression.<sup>30</sup> |
| Access to care | 9 | Assessed in primary and secondary care settings for cardiovascular disease via various prescribing models including dependent, independent, supplementary, and collaborative prescribing,<sup>5, 26, 31, 44, 52</sup> respiratory conditions (dependent and supplementary prescribing),<sup>26, 52</sup> diabetes mellitus (dependent and independent prescribing),<sup>26, 31, 52</sup> mental health,<sup>5, 26, 52</sup> oncology and pain,<sup>26, 45, 52</sup> substance use disorder,<sup>5, 26, 52</sup> general practice,<sup>52</sup> HIV pre-exposure prophylaxis,<sup>5, 54</sup> older adult medicine,<sup>5</sup> and hormonal contraception services.<sup>5, 40, 56</sup> |

CKD: chronic kidney disease

For minor ailments, 14 outcomes were identified. The outcomes evaluated in the most reviews were “Clinical cure or symptom resolution/improvement” and “Cost of service delivery”, both reported in 8 of 14 reviews. For other pharmacist prescribing, 12 outcomes were identified, with “General satisfaction” reported in the most reviews (n=18), followed by “Clinical Effectiveness” and “Resource use due to healthcare utilisation” (both n=15).

## Discussion

### Summary of main findings

This rapid overview synthesised evidence from 43 reviews (systematic, scoping and overviews of reviews) to identify outcomes used to evaluate pharmacist prescribing for minor ailments and other broader clinical contexts from international settings. Across the 14 reviews covering pharmacist prescribing for minor ailments, 14 outcomes were reported, while from 35 reviews in other pharmacist prescribing contexts, 12 outcomes were reported. These identified outcomes broadly encompass clinical, drug-related and prescribing, patient-reported experiences, and economic dimensions, reflecting a comprehensive framework for assessing pharmacist prescribing services.^1, 2^ Several outcomes were frequently cited for evaluating pharmacist prescribing, regardless of the prescribing or clinical context. These include outcomes relevant to clinical improvement, patient experience, cost of care, access to care and guidelines’ concordance/appropriateness of medications. However, some outcomes, such as mortality, prescribing patterns, resource use and GP workload, were context-dependent and more frequently reported in specific clinical or prescribing settings.

Symptom improvement or clinical cure was the most frequently reported clinical outcome of pharmacist prescribing for minor ailments. Similarly, clinical effectiveness and improvements in chronic disease (e.g. blood pressure and glycaemic control, attainment of therapeutic goals) were among the most frequently reported clinical outcomes in evaluating pharmacist prescribing for other broader clinical contexts (e.g. cardiovascular disease, diabetes, mental health). Notably, mortality was cited in identified reviews as an outcome for evaluating the impact of pharmacist prescribing in contexts beyond minor ailments, underscoring the potential for pharmacist prescribing to impact complex, multifactorial and clinically significant endpoints.

Adherence to guidelines and appropriateness of medication were commonly synthesised outcomes in reviews evaluating pharmacist prescribing in minor ailments and other contexts. A key finding in identified reviews evaluating prescribing in the broader context beyond minor ailments was the inclusion of detailed prescribing patterns, which encompassed prescribing rates and medication changes/discontinuations. Evidence suggested that pharmacist prescribing was associated with increased initiation of evidence-based therapies in eligible patients (e.g. antihypertensives, statins, aspirin, contraceptives), more frequent dose changes, and improved deprescribing practices.^5, 30^ In mental health, pharmacist prescribing improved prescribing patterns, most commonly reducing the dosage and absolute number of psychotropic drugs.^38^ This emphasises pharmacists’ role in optimising pharmacotherapy, potentially reducing medication-related harms.

Patient-reported outcomes such as patient experience and satisfaction and access to the care were frequently reported across reviews evaluating the impact of pharmacist prescribing in minor ailments and other contexts across primary and secondary care settings. These outcomes were extensively explored, addressing various aspects, including access to medicines, quality and comprehensiveness of the service, professional relationship, interpersonal communication, continuity and coordination, trust in healthcare providers, patient-reported impacts of care and patient–knowledge and adherence to prescribed medicines.^26, 52, 58^ The ease of accessing a pharmacist, avoiding delays associated with physician appointments, and the convenience of receiving care when medical clinics were unavailable or when patients lacked a regular physician were among the common reasons for choosing a pharmacist prescriber for management of minor ailments and other conditions.^5^ This underscores the potential impact of adopting pharmacist prescribing, not only on patient outcomes but also on more efficient use of healthcare resources.

### Implications and future directions

The identification and synthesis of outcomes across multiple domains demonstrate the variety of potential impacts of pharmacist prescribing, and the findings could support consistent evaluation for pharmacist prescribing. Future studies evaluating pharmacist prescribing, or reviews of such studies, should consider aligning their outcome measurement and synthesis with this structure. However, further work to refine definitions and outcome measures to capture these outcomes would be beneficial. The identified outcomes could inform core outcome set development, and have already been used within a Delphi study to identify critical outcomes for pharmacist prescribing in Ireland.

### Strengths and limitations

The use of an overview of reviews is appropriate to summarise the extent of previous synthesised evidence on a topic, and given the availability of more than 40 systematically conducted reviews which included studies evaluating the effect of pharmacist prescribing. Although it employed many features of a standard overview of reviews, including structured searches of multiple databases and screening and full-text review by two independent reviewers, data extraction was conducted by one reviewer and critical appraisal of included reviews was not conducted (as this was less relevant given the focus of the research question). In addition, by focusing on systematic and scoping reviews, we were unable to include primary studies published since reviews were conducted which may have added further outcomes. However, any outcomes that were not identified in the high number of reviews included would be unlikely to be critically important. Additionally, primary study overlap across the reviews was not quantified. While critical appraisal and overlap identification are important for ensuring reliable conclusions, their omission aligns with circumstances where rapid review methods are tailored to the timeframe and available resources, particularly when conducted for policymakers.^18^ Similar to other reviews, we were unable to clearly define the model of prescribing in some reviews concerning minor ailments, as many included reviews frequently included such conditions but did not clearly delineate the role or scope of independent pharmacist prescribers.^2^

## Conclusions

This rapid overview identified the most frequently synthesised outcome domains for evaluating pharmacist prescribing in minor ailments and other contexts. The identified outcomes encompass clinical, drug-related and prescribing, patient-reported experiences, and economic and other related dimensions. The significance of specific outcomes within these domains are contextually dependent and reflect the acuity of minor ailments versus the chronicity and complexity of managing other conditions. Future research should incorporate core outcome sets to prioritise and standardise outcome measures and definitions, and to enable more robust and setting-specific conclusions on the significant outcomes that appropriately capture the value of pharmacist prescribing across diverse healthcare landscapes.

## Data Availability

All data produced in the present study are available upon reasonable request to the authors

