## Supplementary material for "Identifying outcomes for evaluating the impact of pharmacist prescribing: A rapid overview of reviews": Supplementary File.docx

### Appendix 1 Search strategy

PubMed Search strategy: 29^th^ November 2024

|  | Key Principles | Search terms | Results |
| --- | --- | --- | --- |
| #1 | Pharmacist and Pharmacy | pharmacist[Title/Abstract] OR pharmacy[Title/Abstract] OR "Pharmacists"[Mesh] OR "Pharmacy"[Mesh] | 84,827 |
| #2 | Independent Prescribing | ("independent"[Title/Abstract] OR "autonomous"[Title/Abstract] OR "autonomy"[Title/Abstract] OR "authori*"[Title/Abstract] OR "supplementary"[Title/Abstract] OR "collaborat*"[Title/Abstract] OR "right*"[Title/Abstract] OR "responsibility*"[Title/Abstract]) AND ("prescrib*"[Title/Abstract] OR "prescript*"[Title/Abstract] OR (("exten*"[Title/Abstract] OR "initiat*"[Title/Abstract] OR "substitut*"[Title/Abstract] OR "change*"[Title/Abstract] OR "adjust*"[Title/Abstract]) AND ("medicine*"[Title/Abstract] OR "medication*"[Title/Abstract] OR "prescription*"[Title/Abstract] OR "drug*"[Title/Abstract] OR "Drug Prescriptions"[MeSH Terms] OR "Prescription Drugs"[MeSH Terms] OR "Pharmaceutical Preparations"[MeSH Terms] OR "dose"[Title/Abstract] OR "dosage"[Title/Abstract] OR "Deprescriptions"[MeSH Terms]))) | 118,518 |
| #3 | Pharmacy Prescribing | "pharmacist prescribing"[Title/Abstract:~2] OR "pharmacist prescription"[Title/Abstract:~2] OR "pharmacist prescriptions"[Title/Abstract:~2] OR "pharmacist prescribed"[Title/Abstract:~2] OR "non-medical prescribing"[Title/Abstract:~2] OR "Drug Substitution"[MeSH Terms] OR "non medical switch*"[Title/Abstract] OR (("exten*"[Title/Abstract] OR "modif*"[Title/Abstract] OR "adjust*"[Title/Abstract]) AND ("prescrib*"[Title/Abstract] OR "prescript*"[Title/Abstract])) | 61,718 |
| #4 | Management/Prescribing for Common Conditions | ("Common"[Title/Abstract] OR "self-limiting"[Title/Abstract] OR "minor"[Title/Abstract]) AND ("condition*"[Title/Abstract] OR "illness*"[Title/Abstract] OR "ailment*"[Title/Abstract]) AND ("prescrib*"[Title/Abstract] OR "treat*"[Title/Abstract] OR "manag*"[Title/Abstract] OR "care"[Title/Abstract] OR "service"[Title/Abstract] OR "supply"[Title/Abstract] OR "Therapeutics"[MeSH Terms] OR "Pharmaceutical Services"[MeSH Terms] OR "Disease Management"[MeSH Terms]) | 163,447 |
| #5 | Reviews filtering | ("Systematic Review"[Publication Type:NoExp] OR "Systematic Reviews as Topic"[mesh:noexp] OR "Cochrane Database Syst Rev"[Journal] OR "Evid Rep Technol Assess (Full Rep)"[jour] OR "Evid Rep Technol Assess (Summ)"[jour] OR "scoping"[TI] OR "systematic"[TI] OR ((("comprehensive analysis" [TIAB:~1] OR "comprehensive review" [TIAB:~1] OR "comprehensively reviewed" [TIAB:~1] OR "literature search" [TIAB:~1] OR "literature searches" [TIAB:~1] OR "scoping search" [TIAB:~1] OR "scoping searches" [TIAB:~1]) NOT "narrative review"[TI]) OR "pooled study" [TIAB:~1] OR "systematic search" [TIAB:~1] OR "systematic searches" [TIAB:~1] OR "systematically searched" [TIAB:~1) AND (databases[TIAB] OR "cinahl" [TIAB] OR "cochrane" [TIAB] OR "embase" [TIAB] OR "psycinfo" [TIAB] OR "pubmed" [TIAB] OR "medline" [TIAB] OR "scopus" [TIAB] OR "web science" [TIAB:~1] OR "bibliographic review" [TIAB:~1] OR "bibliographic reviews" [TIAB:~1] OR "literature review" [TIAB:~1] OR "literature reviews" [TIAB:~1]) OR (("electronic database" [TIAB:~1] OR "electronic databases" [TIAB:~1] OR "databases searched" [TIAB:~3]) AND (eligibility [TIAB] OR excluded [TIAB] OR exclusion [TIAB] OR included [TIAB] OR inclusion [TIAB])) OR ("comparative effectiveness" [TIAB:~1] AND "effectiveness review" [TIAB:~2]) OR ("critical interpretive" [TIAB:~1] AND ("interpretive review" [TIAB:~0] OR "interpretive synthesis" [TIAB:~0])) OR ("diagnostic test" [TIAB:~0] AND ("accuracy review" [TIAB] OR "accuracy reviews" [TIAB] OR "accuracy studies" [TIAB] OR "accuracy study" [TIAB]) AND (meta-analysis [TIAB] OR scoping [TIAB] OR systematic [TIAB])) OR ("evidence assessment" [TIAB] AND GRADE [TIAB]) OR ("evidence gap" [TIAB:~2] AND "gap map" [TIAB:~0]) OR "evidence mapping" [TIAB] OR "evidence review" [TIAB] OR "exploratory review" [TIAB] OR "framework synthesis" [TIAB] OR "mapping review" [TIAB:~1] OR "meta epidemiological" [TIAB] OR "meta ethnographic" [TIAB:~0] OR metaethnographic [TIAB] OR "meta ethnography" [TIAB:~0] OR metaethnography [TIAB] OR "meta interpretation" [TIAB:~1] OR "meta narrative" [TIAB:~1] OR "meta review" [TIAB:~1] OR "meta study" [TIAB:~1] OR "meta synthesis" [TIAB:~0] OR metasynthesis [TIAB] OR "meta summary" [TIAB:~1] OR "meta theory" [TIAB:~1] OR "methodological review" [TIAB:~1] OR "methodology review" [TIAB:~1] OR ("mixed methods" [TIAB:~0] AND "methods review" [TIAB:~1]) OR ("mixed methods" [TIAB:~0] AND "methods synthesis" [TIAB:~1]) OR "narrative synthesis" [TIAB:~1] OR "overview reviews" [TIAB:~4] OR ("PRISMA" [TIAB] AND (guideline [TIAB] OR guidelines [TIAB] OR preferred [TIAB] OR reporting [TIAB] OR requirements [TIAB])) OR "PRISMA-P" [TIAB:~0] OR "prognostic review" [TIAB:~1] OR "psychometric review" [TIAB:~1] OR ("qualitative evidence" [TIAB:~0] AND "evidence synthesis" [TIAB:~0]) OR ("qualitative research" [TIAB:~0] AND "research synthesis" [TIAB:~0]) OR ("rapid evidence" [TIAB:~0] AND "evidence assessment" [TIAB:~0]) OR "rapid realist" [TIAB:~0] OR "rapid review" [TIAB:~1] OR "rapid reviews" [TIAB:~1] OR "realist review" [TIAB:~1] OR ("review economic" [TIAB:~1] AND ("economic evaluation" [TIAB:~1] OR "economic evaluations" [TIAB:~1])) OR "review reviews" [TIAB:~1] OR "realist syntheses" [TIAB:~1] OR "realist synthesis" [TIAB:~1] OR "scoping review" [TIAB:~2] OR "scoping reviews" [TIAB:~2] OR "scoping studies" [TIAB:~2] OR "scoping study" [TIAB:~2] OR "systematic evidence map" [TIAB] OR "systematic mapping" [TIAB:~2] OR "systematic literature" [TIAB:~1] OR "systematic Medline" [TIAB:~2] OR "systematic PubMed" [TIAB:~2] OR "systematic review" [TIAB:~2] OR "systematic reviews" [TIAB:~2] OR "systematical review" [TIAB:~1] OR "systematical reviews" [TIAB:~2] OR "systematically identified" [TIAB:~1] OR "systematically review" [TIAB:~1] OR "systematically reviewed" [TIAB:~1] OR "systematized review" [TIAB:~1] OR "umbrella review" [TIAB:~2] OR "umbrella reviews" [TIAB:~2]) | 514,711 |
| #6 | #1 AND (#2 OR #3 OR #4) AND #5 | ("pharmacist"[Title/Abstract] OR "Pharmacy"[Title/Abstract] OR "Pharmacists"[MeSH Terms] OR "Pharmacy"[MeSH Terms]) AND ((("independent"[Title/Abstract] OR "autonomous"[Title/Abstract] OR "autonomy"[Title/Abstract] OR "authori*"[Title/Abstract] OR "supplementary"[Title/Abstract] OR "collaborat*"[Title/Abstract] OR "right*"[Title/Abstract] OR "responsibility*"[Title/Abstract]) AND ("prescrib*"[Title/Abstract] OR "prescript*"[Title/Abstract] OR (("exten*"[Title/Abstract] OR "initiat*"[Title/Abstract] OR "substitut*"[Title/Abstract] OR "change*"[Title/Abstract] OR "adjust*"[Title/Abstract]) AND ("medicine*"[Title/Abstract] OR "medication*"[Title/Abstract] OR "prescription*"[Title/Abstract] OR "drug*"[Title/Abstract] OR "Drug Prescriptions"[MeSH Terms] OR "Prescription Drugs"[MeSH Terms] OR "Pharmaceutical Preparations"[MeSH Terms] OR "dose"[Title/Abstract] OR "dosage"[Title/Abstract] OR "Deprescriptions"[MeSH Terms])))) OR ("pharmacist prescribing"[Title/Abstract:~2] OR "pharmacist prescription"[Title/Abstract:~2] OR "pharmacist prescriptions"[Title/Abstract:~2] OR "pharmacist prescribed"[Title/Abstract:~2] OR "non-medical prescribing"[Title/Abstract:~2] OR "Drug Substitution"[MeSH Terms] OR "non medical switch*"[Title/Abstract] OR (("exten*"[Title/Abstract] OR "modif*"[Title/Abstract] OR "adjust*"[Title/Abstract]) AND ("prescrib*"[Title/Abstract] OR "prescript*"[Title/Abstract]))) OR (("Common"[Title/Abstract] OR "self-limiting"[Title/Abstract] OR "minor"[Title/Abstract]) AND ("condition*"[Title/Abstract] OR "illness*"[Title/Abstract] OR "ailment*"[Title/Abstract]) AND ("prescrib*"[Title/Abstract] OR "treat*"[Title/Abstract] OR "manag*"[Title/Abstract] OR "care"[Title/Abstract] OR "service"[Title/Abstract] OR "supply"[Title/Abstract] OR "Therapeutics"[MeSH Terms] OR "Pharmaceutical Services"[MeSH Terms] OR "Disease Management"[MeSH Terms]))) AND ((("Systematic Review"[Publication Type:noexp] OR "Systematic Reviews as Topic"[MeSH Terms:noexp] OR "Cochrane Database Syst Rev"[Journal] OR "evid rep technol assess full rep"[Journal] OR "evid rep technol assess summ"[Journal] OR "scoping"[Title] OR "systematic"[Title] OR ((("comprehensive analysis"[Title/Abstract:~1] OR "comprehensive review"[Title/Abstract:~1] OR "comprehensively reviewed"[Title/Abstract:~1] OR "literature search"[Title/Abstract:~1] OR "literature searches"[Title/Abstract:~1] OR "scoping search"[Title/Abstract:~1] OR "scoping searches"[Title/Abstract:~1]) NOT "narrative review"[Title]) OR "pooled study"[Title/Abstract:~1] OR "systematic search"[Title/Abstract:~1] OR "systematic searches"[Title/Abstract:~1] OR "systematically searched"[Title/Abstract:~1])) AND ("databases"[Title/Abstract] OR "cinahl"[Title/Abstract] OR "cochrane"[Title/Abstract] OR "embase"[Title/Abstract] OR "psycinfo"[Title/Abstract] OR "pubmed"[Title/Abstract] OR "medline"[Title/Abstract] OR "scopus"[Title/Abstract] OR "web science"[Title/Abstract:~1] OR "bibliographic review"[Title/Abstract:~1] OR "bibliographic reviews"[Title/Abstract:~1] OR "literature review"[Title/Abstract:~1] OR "literature reviews"[Title/Abstract:~1])) OR (("electronic database"[Title/Abstract:~1] OR "electronic databases"[Title/Abstract:~1] OR "databases searched"[Title/Abstract:~3]) AND ("eligibility"[Title/Abstract] OR "excluded"[Title/Abstract] OR "exclusion"[Title/Abstract] OR "included"[Title/Abstract] OR "inclusion"[Title/Abstract])) OR ("comparative effectiveness"[Title/Abstract:~1] AND "effectiveness review"[Title/Abstract:~2]) OR ("critical interpretive"[Title/Abstract:~1] AND ("interpretive review"[Title/Abstract:~0] OR "interpretive synthesis"[Title/Abstract:~0])) OR ("diagnostic test"[Title/Abstract:~0] AND ("accuracy review"[Title/Abstract] OR "accuracy reviews"[Title/Abstract] OR "accuracy studies"[Title/Abstract] OR "accuracy study"[Title/Abstract]) AND ("meta-analysis"[Title/Abstract] OR "scoping"[Title/Abstract] OR "systematic"[Title/Abstract])) OR ("evidence assessment"[Title/Abstract] AND "GRADE"[Title/Abstract]) OR ("evidence gap"[Title/Abstract:~2] AND "gap map"[Title/Abstract:~0]) OR "evidence mapping"[Title/Abstract] OR "evidence review"[Title/Abstract] OR "exploratory review"[Title/Abstract] OR "framework synthesis"[Title/Abstract] OR "mapping review"[Title/Abstract:~1] OR "meta epidemiological"[Title/Abstract] OR "meta ethnographic"[Title/Abstract:~0] OR "metaethnographic"[Title/Abstract] OR "meta ethnography"[Title/Abstract:~0] OR "metaethnography"[Title/Abstract] OR "meta interpretation"[Title/Abstract:~1] OR "meta narrative"[Title/Abstract:~1] OR "meta review"[Title/Abstract:~1] OR "meta study"[Title/Abstract:~1] OR "meta synthesis"[Title/Abstract:~0] OR "metasynthesis"[Title/Abstract] OR "meta summary"[Title/Abstract:~1] OR "meta theory"[Title/Abstract:~1] OR "methodological review"[Title/Abstract:~1] OR "methodology review"[Title/Abstract:~1] OR ("mixed methods"[Title/Abstract:~0] AND "methods review"[Title/Abstract:~1]) OR ("mixed methods"[Title/Abstract:~0] AND "methods synthesis"[Title/Abstract:~1]) OR "narrative synthesis"[Title/Abstract:~1] OR "overview reviews"[Title/Abstract:~4] OR ("PRISMA"[Title/Abstract] AND ("guideline"[Title/Abstract] OR "guidelines"[Title/Abstract] OR "preferred"[Title/Abstract] OR "reporting"[Title/Abstract] OR "requirements"[Title/Abstract])) OR "PRISMA-P"[Title/Abstract:~0] OR "prognostic review"[Title/Abstract:~1] OR "psychometric review"[Title/Abstract:~1] OR ("qualitative evidence"[Title/Abstract:~0] AND "evidence synthesis"[Title/Abstract:~0]) OR ("qualitative research"[Title/Abstract:~0] AND "research synthesis"[Title/Abstract:~0]) OR ("rapid evidence"[Title/Abstract:~0] AND "evidence assessment"[Title/Abstract:~0]) OR "rapid realist"[Title/Abstract:~0] OR "rapid review"[Title/Abstract:~1] OR "rapid reviews"[Title/Abstract:~1] OR "realist review"[Title/Abstract:~1] OR ("review economic"[Title/Abstract:~1] AND ("economic evaluation"[Title/Abstract:~1] OR "economic evaluations"[Title/Abstract:~1])) OR "review reviews"[Title/Abstract:~1] OR "realist syntheses"[Title/Abstract:~1] OR "realist synthesis"[Title/Abstract:~1] OR "scoping review"[Title/Abstract:~2] OR "scoping reviews"[Title/Abstract:~2] OR "scoping studies"[Title/Abstract:~2] OR "scoping study"[Title/Abstract:~2] OR "systematic evidence map"[Title/Abstract] OR "systematic mapping"[Title/Abstract:~2] OR "systematic literature"[Title/Abstract:~1] OR "systematic Medline"[Title/Abstract:~2] OR "systematic PubMed"[Title/Abstract:~2] OR "Systematic Review"[Title/Abstract:~2] OR "systematic reviews"[Title/Abstract:~2] OR "systematical review"[Title/Abstract:~1] OR "systematical reviews"[Title/Abstract:~2] OR "systematically identified"[Title/Abstract:~1] OR "systematically review"[Title/Abstract:~1] OR "systematically reviewed"[Title/Abstract:~1] OR "systematized review"[Title/Abstract:~1] OR "umbrella review"[Title/Abstract:~2] OR "umbrella reviews"[Title/Abstract:~2]) | 352 |

Embase Search strategy: 29^th^ November 2024

|  | Key Principles | Search terms | Results |
| --- | --- | --- | --- |
| #1 | Pharmacist and Pharmacy | 'pharmacist':ti,ab,kw OR 'pharmacist'/exp OR 'pharmacy (shop)'/exp OR 'pharmacy':ti,ab,kw | 230,585 |
| #2 | Independent Prescribing | (independent:ti,ab,kw OR autonomous:ti,ab,kw OR autonomy:ti,ab,kw OR authori*:ti,ab,kw OR supplementary:ti,ab,kw OR collaborat*:ti,ab,kw OR right*:ti,ab,kw OR responsibility*:ti,ab,kw) AND (prescrib*:ti,ab,kw OR prescript*:ti,ab,kw OR ((exten*:ti,ab,kw OR initiat*:ti,ab,kw OR substitut*:ti,ab,kw OR change*:ti,ab,kw OR adjust*:ti,ab,kw) AND (medicine*:ti,ab,kw OR medication*:ti,ab,kw OR prescription*:ti,ab,kw OR drug*:ti,ab,kw OR dose:ti,ab,kw OR dosage:ti,ab,kw OR 'prescription'/exp OR 'prescription drug'/exp OR 'deprescription'/exp))) | 266,024 |
| #3 | Pharmacy Prescribing | ((pharmacist NEAR/2 prescribing):ab) OR ((pharmacist NEAR/2 prescription):ab) OR ((pharmacist NEAR/2 prescriptions):ab) OR ((pharmacist NEAR/2 prescribed):ab) OR (('non medical' NEAR/2 prescribing):ab) OR 'drug substitution'/exp OR ('non-medical':ti,ab,kw AND switch*:ti,ab,kw) OR ((exten*:ti,ab,kw OR modif*:ti,ab,kw OR adjust*:ti,ab,kw) AND (prescrib*:ti,ab,kw OR prescript*:ti,ab,kw)) | 157,439 |
| #4 | Management/Prescribing for Common Conditions | (common:ti,ab,kw OR 'self limiting':ti,ab,kw OR minor:ti,ab,kw) AND (condition*:ti,ab,kw OR illness*:ti,ab,kw OR ailment*:ti,ab,kw) AND (prescrib*:ti,ab,kw OR treat*:ti,ab,kw OR manag*:ti,ab,kw OR care:ti,ab,kw OR service:ti,ab,kw OR supply:ti,ab,kw OR 'therapy'/exp OR 'disease management'/exp) | 283,707 |
| #5 | Reviews filtering | 'systematic review'/de OR 'systematic review (topic)'/de OR (('comprehensive':ti,ab,kw OR 'mapping':ti,ab,kw OR 'methodology':ti,ab,kw OR 'scoping':ti,ab,kw OR 'systematic':ti,ab,kw) AND ('search':ti,ab,kw OR 'searched':ti,ab,kw OR 'searches':ti,ab,kw OR 'studies':ti,ab,kw) AND ('cinahl':ti,ab,kw OR 'cochrane':ti,ab,kw OR 'embase':ti,ab,kw OR 'psycinfo':ti,ab,kw OR 'pubmed':ti,ab,kw OR 'medline':ti,ab,kw OR 'scopus':ti,ab,kw OR 'web of science':ti,ab,kw OR 'bibliographic review':ti,ab,kw OR 'bibliographic reviews':ti,ab,kw OR 'literature review':ti,ab,kw OR 'literature reviews':ti,ab,kw OR 'literature search':ti,ab,kw OR 'literature searches':ti,ab,kw OR 'qualitative review':ti,ab,kw OR 'qualitative reviews':ti,ab,kw OR 'quantitative review':ti,ab,kw OR 'quantitative reviews':ti,ab,kw)) OR 'comprehensive review':ti,ab,kw OR 'comprehensive reviews':ti,ab,kw OR 'comprehensive search':ti,ab,kw OR 'comprehensive searches':ti,ab,kw OR 'critical review':ti,ab,kw OR 'critical reviews':ti,ab,kw OR (('electronic database':ti,ab,kw OR 'electronic databases':ti,ab,kw OR (databases NEAR/3 searched)) AND (eligibility:ti,ab,kw OR excluded:ti,ab,kw OR exclusion:ti,ab,kw OR included:ti,ab,kw OR inclusion:ti,ab,kw)) OR 'evidence assessment':ti,ab,kw OR 'evidence review':ti,ab,kw OR 'exploratory review':ti,ab,kw OR 'framework synthesis':ti,ab,kw OR 'mapping review':ti,ab,kw OR 'meta-review':ti,ab,kw OR 'meta-synthesis':ti,ab,kw OR 'methodology review':ti,ab,kw OR 'mixed methods review':ti,ab,kw OR 'mixed methods synthesis':ti,ab,kw OR (overview NEAR/4 reviews) OR 'prisma':ab OR ('preferred':ti,ab,kw AND reporting:ti,ab,kw) OR 'prognostic review':ti,ab,kw OR 'psychometric review':ti,ab,kw OR 'rapid evidence assessment':ti,ab,kw OR 'rapid literature review':ti,ab,kw OR 'rapid literature search':ti,ab,kw OR 'rapid realist':ti,ab,kw OR 'rapid review':ti,ab,kw OR 'rapid reviews':ti,ab,kw OR 'realist review':ti,ab,kw OR 'review of reviews':ti,ab,kw OR 'scoping review':ti,ab,kw OR 'scoping reviews':ti,ab,kw OR 'scoping study':ti,ab,kw OR 'systematic evidence map':ti,ab,kw OR 'systematic evidence mapping':ti,ab,kw OR 'systematic literature':ti,ab,kw OR 'systematic medline':ti,ab,kw OR 'systematic pubmed':ti,ab,kw OR 'systematic review':ti,ab,kw OR 'systematic reviews':ti,ab,kw OR 'systematic search':ti,ab,kw OR 'systematic searches':ti,ab,kw OR 'systematical literature review':ti,ab,kw OR 'systematical review':ti,ab,kw OR 'systematical reviews':ti,ab,kw OR 'systematically identified':ti,ab,kw OR 'systematically review':ti,ab,kw OR 'systematically reviewed':ti,ab,kw OR 'umbrella review':ti,ab,kw OR 'umbrella reviews':ti,ab,kw OR '13616137':is OR 'cochrane database of systematic reviews'/jt | 780058 |
| #6 | #1 AND (#2 OR #3 OR #4) AND #5 | ('pharmacist':ti,ab,kw OR 'pharmacist'/exp OR 'pharmacy (shop)'/exp OR 'pharmacy':ti,ab,kw) AND ((independent:ti,ab,kw OR autonomous:ti,ab,kw OR autonomy:ti,ab,kw OR authori*:ti,ab,kw OR supplementary:ti,ab,kw OR collaborat*:ti,ab,kw OR right*:ti,ab,kw OR responsibility*:ti,ab,kw) AND (prescrib*:ti,ab,kw OR prescript*:ti,ab,kw OR ((exten*:ti,ab,kw OR initiat*:ti,ab,kw OR substitut*:ti,ab,kw OR change*:ti,ab,kw OR adjust*:ti,ab,kw) AND (medicine*:ti,ab,kw OR medication*:ti,ab,kw OR prescription*:ti,ab,kw OR drug*:ti,ab,kw OR dose:ti,ab,kw OR dosage:ti,ab,kw OR 'prescription'/exp OR 'prescription drug'/exp OR 'deprescription'/exp))) OR ((pharmacist NEAR/2 prescribing):ab) OR ((pharmacist NEAR/2 prescription):ab) OR ((pharmacist NEAR/2 prescriptions):ab) OR ((pharmacist NEAR/2 prescribed):ab) OR (('non medical' NEAR/2 prescribing):ab) OR 'drug substitution'/exp OR ('non-medical':ti,ab,kw AND switch*:ti,ab,kw) OR ((exten*:ti,ab,kw OR modif*:ti,ab,kw OR adjust*:ti,ab,kw) AND (prescrib*:ti,ab,kw OR prescript*:ti,ab,kw)) OR ((common:ti,ab,kw OR 'self limiting':ti,ab,kw OR minor:ti,ab,kw) AND (condition*:ti,ab,kw OR illness*:ti,ab,kw OR ailment*:ti,ab,kw) AND (prescrib*:ti,ab,kw OR treat*:ti,ab,kw OR manag*:ti,ab,kw OR care:ti,ab,kw OR service:ti,ab,kw OR supply:ti,ab,kw OR 'therapy'/exp OR 'disease management'/exp))) AND ('systematic review'/de OR 'systematic review (topic)'/de OR (('comprehensive':ti,ab,kw OR 'mapping':ti,ab,kw OR 'methodology':ti,ab,kw OR 'scoping':ti,ab,kw OR 'systematic':ti,ab,kw) AND ('search':ti,ab,kw OR 'searched':ti,ab,kw OR 'searches':ti,ab,kw OR 'studies':ti,ab,kw) AND ('cinahl':ti,ab,kw OR 'cochrane':ti,ab,kw OR 'embase':ti,ab,kw OR 'psycinfo':ti,ab,kw OR 'pubmed':ti,ab,kw OR 'medline':ti,ab,kw OR 'scopus':ti,ab,kw OR 'web of science':ti,ab,kw OR 'bibliographic review':ti,ab,kw OR 'bibliographic reviews':ti,ab,kw OR 'literature review':ti,ab,kw OR 'literature reviews':ti,ab,kw OR 'literature search':ti,ab,kw OR 'literature searches':ti,ab,kw OR 'qualitative review':ti,ab,kw OR 'qualitative reviews':ti,ab,kw OR 'quantitative review':ti,ab,kw OR 'quantitative reviews':ti,ab,kw)) OR 'comprehensive review':ti,ab,kw OR 'comprehensive reviews':ti,ab,kw OR 'comprehensive search':ti,ab,kw OR 'comprehensive searches':ti,ab,kw OR 'critical review':ti,ab,kw OR 'critical reviews':ti,ab,kw OR (('electronic database':ti,ab,kw OR 'electronic databases':ti,ab,kw OR (databases NEAR/3 searched)) AND (eligibility:ti,ab,kw OR excluded:ti,ab,kw OR exclusion:ti,ab,kw OR included:ti,ab,kw OR inclusion:ti,ab,kw)) OR 'evidence assessment':ti,ab,kw OR 'evidence review':ti,ab,kw OR 'exploratory review':ti,ab,kw OR 'framework synthesis':ti,ab,kw OR 'mapping review':ti,ab,kw OR 'meta-review':ti,ab,kw OR 'meta-synthesis':ti,ab,kw OR 'methodology review':ti,ab,kw OR 'mixed methods review':ti,ab,kw OR 'mixed methods synthesis':ti,ab,kw OR (overview NEAR/4 reviews) OR 'prisma':ab OR ('preferred':ti,ab,kw AND reporting:ti,ab,kw) OR 'prognostic review':ti,ab,kw OR 'psychometric review':ti,ab,kw OR 'rapid evidence assessment':ti,ab,kw OR 'rapid literature review':ti,ab,kw OR 'rapid literature search':ti,ab,kw OR 'rapid realist':ti,ab,kw OR 'rapid review':ti,ab,kw OR 'rapid reviews':ti,ab,kw OR 'realist review':ti,ab,kw OR 'review of reviews':ti,ab,kw OR 'scoping review':ti,ab,kw OR 'scoping reviews':ti,ab,kw OR 'scoping study':ti,ab,kw OR 'systematic evidence map':ti,ab,kw OR 'systematic evidence mapping':ti,ab,kw OR 'systematic literature':ti,ab,kw OR 'systematic medline':ti,ab,kw OR 'systematic pubmed':ti,ab,kw OR 'systematic review':ti,ab,kw OR 'systematic reviews':ti,ab,kw OR 'systematic search':ti,ab,kw OR 'systematic searches':ti,ab,kw OR 'systematical literature review':ti,ab,kw OR 'systematical review':ti,ab,kw OR 'systematical reviews':ti,ab,kw OR 'systematically identified':ti,ab,kw OR 'systematically review':ti,ab,kw OR 'systematically reviewed':ti,ab,kw OR 'umbrella review':ti,ab,kw OR 'umbrella reviews':ti,ab,kw OR '13616137':is OR 'cochrane database of systematic reviews'/jt) | 917 |

Cochrane Database of Systematic Reviews Search Strategy: 29^th^ November 2024

|  | Key Principles | Search terms | Total Results (reviews no) |
| --- | --- | --- | --- |
| #1 | Pharmacist and Pharmacy | (pharmacist* or pharmacy):ti,ab,kw | 10986 |
| #2 |  | MeSH descriptor: [Pharmacists] explode all trees | 1139 |
| #3 |  | MeSH descriptor: [Pharmacies] explode all trees | 224 |
| #4 |  | #1 or #2 or #3 | 11000 (74) |
| #5 | Independent Prescribing | ((medicine* OR medication* OR prescription* OR drug* OR dose OR dosage)):ti,ab,kw | 1030801 |
| #6 |  | MeSH descriptor: [Prescription Drugs] explode all trees | 170 |
| #7 |  | MeSH descriptor: [Deprescriptions] explode all trees | 117 |
| #8 |  | ((exten* OR initiat* OR substitut* OR change* OR adjust*)):ti,ab,kw | 628294 |
| #9 |  | (prescrib* or prescript*):ti,ab,kw | 49749 |
| #10 |  | ((independent or autonomous or autonomy or authori* or supplementary or collaborat* or right* or responsibility*)):ti,ab,kw | 154330 |
| #11 |  | #10 and (((#5 or #6 or #7) and #8) or #9) | 33188 (450) |
| #12 | Pharmacy Prescribing | ((pharmacist NEAR/2 prescribing) or (pharmacist NEAR/2 prescription) or (pharmacist NEAR/2 prescriptions) or (pharmacist NEAR/2 prescribed) or (non-medical NEAR/2 prescribing) or (non-medical NEAR/2 switch)) | 254 |
| #13 |  | MeSH descriptor: [Drug Substitution] explode all trees | 548 |
| #14 |  | ((exten* OR modif* OR adjust*) AND (prescrib* OR prescript*)):ti,ab,kw | 10855 |
| #15 |  | #12 or #13 or #14 | 11581 (159) |
| #16 | Management/Prescribing for Common Conditions | (prescrib* or treat* or manag* or care or service or supply):ti,ab,kw | 1327476 |
| #17 |  | MeSH descriptor: [Therapeutics] explode all trees | 436361 |
| #18 |  | MeSH descriptor: [Disease Management] explode all trees | 7645 |
| #19 |  | ((Common or self-limiting or minor) and (condition* or illness* or ailment*)):ti,ab,kw | 30348 |
| #20 |  | (16 or #17 or #18) and 19 | 109830 (8266) |
| #14 | #4 AND (#11 OR #15 OR #20) |  | 2214 (71 reviews) |

Epistemonikos Search strategy: 29th November 2024

|  | Key Principles | Search terms | Results of Broad Synthesis | Results of Systematic Reviews |
| --- | --- | --- | --- | --- |
| #1 | Pharmacist and Pharmacy | [Title/abstract]: Pharmacist* OR pharmacy | 409 | 2335 |
| #2 | Independent Prescribing | [Title/abstract]: (independent OR autonomous OR autonomy OR authori* OR supplementary OR collaborat* OR right* OR responsibility*) AND (prescrib* OR prescript* OR ((exten* OR initiat* OR substitut* OR change* OR adjust*) AND (medicine* OR medication* OR prescription* OR drug* OR “Prescription Drugs” OR dose OR dosage OR Deprescript* OR Deprescrib*))) | 352 | 3966 |
| #3 | Pharmacy Prescribing | [Title/abstract]: “pharmacist prescribing” OR “pharmacist prescription” OR “pharmacist prescriptions” OR “pharmacist prescribed” OR “non-medical prescribing” OR “non-medical switch*” OR ((exten* OR modif* OR adjust*) AND (prescrib* OR prescript*)) | 165 | 1636 |
| #4 | Management/Prescribing for Common Conditions | [Title/abstract]: (Common OR self-limiting OR minor) AND (condition* OR illness* OR ailment*) AND (prescrib* OR treat* OR manag* OR care OR service OR supply OR Therapeutics) | 525 | 7044 |
| #6 | #1 AND (#2 OR #3 OR #4) |  | 71 | 316 |

### Appendix 2 Characteristics of included reviews

| **Authors** | **Study Design** | **Setting** | **Disease area** | **Type of prescribing** | **Outcome** |
| --- | --- | --- | --- | --- | --- |
| Ali et al. 2024  [1] | Scoping review | Community | Cardiovascular disease and diabetes mellitus | Independent and Dependent prescribing | Medication Adherence |
| Alshammari et al. 2023  [2] | Systematic review | Secondary care | Mental health and aged care | Collaborative and interim prescribers | Clinical outcomes (reduction in emergency visits and patient satisfaction) |
| Ardavani et al. 2024  [3] | Systematic review | Primary care | Chronic kidney disease (CKD) | Collaborative and Independent prescribing | Clinical outcomes (parathyroid hormone (PTH), blood pressure (BP) control, changes in estimated CV risk, lipid management); patient satisfaction |
| Atey et al. 2022  [4] | Systematic review | Secondary care | ED patients | Collaborative/supplementary and Independent | Medication errors |
| Babashahi et al. 2023  [5] | Scoping review | Primary and secondary care | Cardiovascular and chronic pain | Independent | Cost-effectiveness analysis |
| Badran et al. 2024  [6] | Scoping review | Primary | Urinary tract infection (UTI) | Patient Group Directions (PGDs) and minor ailment prescribing | Clinical cure (i.e., symptom resolution), adverse events, medication appropriateness, patient adherence, number of follow-ups, treatment failures (and reasons for), referral rates, time saving, cost, GP workload and patient satisfaction. |
| Bhanbhro et al. 2011  [7] | Systematic review | Primary care | Cardiovascular | Supplementary prescribing | Effectiveness and patients experience (access and acceptability) |
| Buckingham et al. 2021  [8] | Scoping review | Community | Obstetrics and gynaecology | Patient Group Direction/Collaborative prescribing agreement | Clinical (reduction in unintended pregnancy rates), accessibility and cost saving |
| Cao et al.2021  [9] | Systematic review | HF clinics | Heart failure | Collaborative and independent | Clinical (death, all-cause hospitalisation and HF hospitalisation, and medication errors or discrepancies) and appropriateness of medications (the proportion of HFrEF patients taking any GDMT and GDMT at target doses) |
| Curley et al. 2016  [10] | Systematic review | Primary care | Minor ailments (e.g. eye infection, head lice, headache/back pain, dyspepsia) and erectile dysfunction |  | Symptom resolution/improvement, cost, satisfaction, GP consultation/ referral, patient compliance, appropriateness of medications, and change in number of chloramphenicol prescriptions. |
| Dawoud et al. 2019  [11] | Systematic review | Primary care | Chronic pain | Independent prescribing | Cost-effectiveness |
| Famiyeh and McCarthy 2017  [12] | Scoping review | Primary and secondary care | General practice, mental health, cardiovascular | Dependent and Independent | Patient experience |
| Faruquee and Guirguis 2015  [13] | Scoping review | Primary | Minor ailments, cardiovascular, diabetes mellitus and obstetrics/gynaecology | Dependent Per protocol, collaborative prescribing | Clinical (symptom resolution, glycaemic control, BP control, LDL improve, detection of an underlying disease, quality of life, survival rates), cost effectiveness, improved medication use, increased drug-related problem identification |
| Finley, Crismon, and Rush 2023  [14] | Systematic review | Primary and secondary care | Mental health | Dependent (by protocol) | Clinical (improvement); cost; and adherence |
| Greer et al. 2015  [15] | Systematic review | Primary and secondary | Cardiovascular, chronic KidneyDisease | Unclear | All-cause mortality; remission/improved outcomes; BP/ HbA1c/ LDL goals attainment; prescribing outcomes (inappropriate dosage/prescription or omission, non-adherence to prescribed regimen, adverse events); cost; health-related quality of life; access to care; and patient satisfaction |
| Jebara et al. 2018  [16] | Systematic review | Primary care and secondary care | Respiratory, cardiovascular, diabetes, oncology and pain, mental health, smoking cessation, arthritis, and minor ailments | Independent and supplementary prescribing | Patient experience/satisfaction |
| Jordan et al. 2021  [17] | Scoping review | General practice | Opioid Analgesic Dependence | Independent prescribing | Appropriateness of prescription |
| Kc et al. 2022  [18] | Systematic review | Primary care/ ambulatory | Travel medicine | Collaborative and Independent | Accessibility; Patients’ and travellers’ satisfaction |
| Kennedy et al. 2019  [19] | Systematic review | Primary | Obstetrics/gynaecology |  | Patient satisfaction |
| Krstic Devaud, and Sadeghipour 2021  [20] | Systematic review | Primary and secondary | Cardiovascular, diabetes, mental health, urogenital, COPD, and gastroenterology (peptic ulcer and gastroesophageal reflux diseases | Non-medical switching | Cost |
| Leong et al. 2021  [21] | Overview of reviews | Primary and secondary | Minor ailments cardiovascular, chronic pain and other chronic diseases | Independent | Clinical (symptom resolution, BP and glycaemic control), reconsultation rate, accessibility, GP workload, patient Satisfaction and cost |
| Liu et al. 2024  [22] | Systematic review | Nursing homes | Geriatric | Independent | Prescribing outcomes (risk of DBI exposure), patient outcomes (mortality, hospital admission, fall, quality of life, healthcare utilization) |
| Lum et al. 2023  [23] | Scoping review | Primary and Secondary | Diabetes mellitus | Independent/ collaborative and unclear | Prescribing outcomes (changes in use rates of pneumococcal vaccination, statin, aspirin), clinical outcomes (improve in HbA1c, BP, FBG, TChol, LDL, TG, HDL, microalbumin testing and attainment of clinical goals), adverse events, patient-reported outcomes (diabetes knowledge, treatment satisfaction, medication adherence, diabetes distress, quality of life, and self-care), service (e.g. general medicine visits, ED visits, hospitalisations) utilisation andeconomic (direct medical costs, quality adjusted life years, and productivity loss from absenteeism, presenteeism, work impairment, activity impairment) |
| Motlohi et al. 2023  [24] | Systematic review | Primary | Diabetes mellitus | Dependent and Independent | Clinical (glycaemic control, major CV event risk reduction); patient satisfaction |
| Naseralallah et al. 2024  [25] | Systematic review | Secondary care | Perioperative settings | Independent prescribing | Appropriateness of antibiotic choice |
| Newlonet al. 2021  [26] | Systematic review | Community | Obstetrics and gynaecology | Independent prescribing | Accessibility and cost saving |
| Noblet et al. 2018  [27] | Systematic review | Primary secondary care | Chronic pain and perioperative setting | Independent and Supplementary prescribing | Patient reported outcomes (pain, anxiety and depression improvement), appropriateness of prescribing (e.g. the number of medications chartered at an incorrect dose or frequency, the number of missed doses of specific medications post operatively, and appropriateness of VTE prophylaxis prescribing), cost-effectiveness |
| Passey et al. 2021  [28] | Systematic review | Primary and secondary care | Oncology | Collaborative | Patient safety (adverse events, drug-drug interactions, DRPs, and medication errors); patient adherence and/or compliance; patient satisfaction; and time and cost savings |
| Paudyalet al. 2018  [29] | Systematic review | Primary and Secondary | Minor ailments related to digestive, respiratory, | Unclear | Clinical outcomes (symptom resolution/improvement, reconsultation); prescribing outcomes (adherence to treatment, side effects, use of any other treatments,); quality of life; general health status, impact of the illness on sleep and daily activities |
| Paudyalet al. 2013  [30] | Systematic review | Primary care (pharmacy settings) | Minor ailments | Independent prescribing | Health-related outcome (resolution of symptoms, consultation and referral rates); cost-related outcomes (mean cost of consultations); and general practice workload |
| (Perraudin, Bugnon, and Pelletier-Fleury 2016  [31] | Systematic review | Primary | Chronic pain and minor alignments | Independent prescribing | Cost effectiveness and saving |
| Piraux et al. 2024  [32] | Scoping review | Primary care | Minor ailments | Independent prescribing | Efficacy (clinical cure and resolution of symptoms); accessibility; safety (adverse effects); and cost (cost saving per patient/ per year |
| Poh et al. 2018  [33] | Systematic review | Secondary | Cardiovascular (VTE, HTN and peripheral vascular diseases), diabetes mellitus, perioperative setting | Dependent (by protocol), Collaborative, Supplementary prescribing | Clinical outcomes (BP control, glycaemic control, cholesterol control, time to therapeutic INR range, subtherapeutic or supratherapeutic INR and adverse effects such as bleeding events, thromboembolic events and deaths); prescribing Outcomes (appropriateness of prescribing, prescribing errors, medication omissions); and patient-reported outcomes (general satisfaction). |
| Rotta et al. 2015  [34] | Overview of reviews | Primary and secondary care | Diabetes mellitus, Mental health, long term care facilities | Dependent (by protocol) and Collaborative | Clinical outcomes (symptom improvement blood pressure, glycaemic and lipid management); patient satisfaction; adherence; cost; prescribing pattern; and appropriateness of prescription |
| Shresthaet al.2021  [35] | Overview of reviews | Primary and secondary | Musculoskeletal systems (knee, spine, joint, back), neurological system (headache and migraine), cancer and unspecified chronic pain. | Unclear | Clinical outcomes (pain intensity and pain relief, adverse drug reactions and drug-related problems, and physical functioning and mental health); Quality of life; patient satisfaction, and cost |
| Varas-Doval et al. 2021  [36] | Systematic review | Primary care | Hypertension | Independent prescribing | Clinical outcomes (change in systolic and diastolic BP at 6 months) |
| Walpola et al. 2024  [37] | Systematic review | Primary and secondary care | Cardiovascular, respiratory, mental health, minor ailments (e.g. group A streptococcal pharyngitis, impetigo, urinary tract infection), HIV pre-exposure prophylaxis, smoking cessation, hormonal contraception and opioid use disorders | Dependent, Collaborative, Independent and unclear | Access to medicines (perceived ease of medicines access, time to medicines access, number of prescriptions or medicines dispensed); and patient-reported outcomes (Perceived ease and convenience, cost and affordability) |
| Weeks et al. 2016  [38] | Systematic review | Primary and Secondary | Ischaemic stroke or transient ischaemic attack, cardiovascular (e.g. hypertension, dyslipidaemia, receiving anticoagulation therapy), diabetes mellitus, depression, chronic pain, aged care and postoperative care. | Supplementary or Independent | Mortality; effectiveness (anticoagulation dose differences, time to therapeutic levels, partial thromboplastin time (sec), improvements in BP, glycaemic and lipid profiles, clinical and functional severity, and adverse events); safety and adverse effects (hospitalisations, emergency department visits, urgent care, same day medical visits for BP problems, hypotension, fainting, loss of consciousness and allergic reactions); prescribing outcomes (doses missed, incorrect doses, incorrect frequencies, medication started, doses changed, drug changed, aspirin and cholesterol-lowering medications started); prescriber adherence to practice guidelines; cost; resource utilisation; patient adherence/compliance; patient satisfaction; quality of life; disability, overall self-rated health, overall rating of health satisfaction and physical activity |
| Wu et al. 2021  [39] | Systematic review | Primary | Acute pharyngitis or sore throat, acute otitis media, acute bacterial sinusitis, chronic bacterial sinusitis, chronic obstructive pulmonary disease (COPD) exacerbation, uncomplicated urinary tract infection (UTI) or cystitis, and cold sores | Patient Group Directions (PGDs) and Independent prescribing | Prescribing outcomes (prescribing rate/antimicrobial use, guideline concordance, drug selection); patient-centred outcomes (Clinical cure/improvement, Treatment failure, Health care utilisation, ADE, Access to care, Patient satisfaction); cost; number of service claims; rate/results of urine dipstick testing, and % of positive Group A Streptococcus tests |
| Wubben and Vivian 2008  [40] | Systematic review | Primary care clinics | Diabetes mellitus | Collaborative | Clinical outcomes (blood pressure, glycaemic and lipid management) and patient satisfaction |
| Yuan et al. 2019  [41] | Systematic review | Primary care | Cardiovascular | Unclear | Effectiveness (BP control, proportion of patients achieving target LDL-c levels) |
| Yusuff, Makhlouf, and Ibrahim 2021  [42] | Systematic review | Primary care | Minor ailments (musculoskeletal pain, cough, colds, eye irritation, allergic rhinitis, dyspepsia, constipation, diarrhoea, ear problems, helminth) | Unclear | Symptoms resolution andcost |
| Zhao et al. 2021  [43] | Scoping review | Primary care | HIV Pre-exposure Prophylaxis | Collaborative | Accessibility; and patient experience |
| ADE: adverse drug event, CKD: chronic kidney disease, COPD: chronic obstructive pulmonary disease, CV: cardiovascular, DBI: drug burden index, ED: emergency department, FBG: fasting blood glucose, GMDT: guideline directed medical therapy, HbA1c: haemoglobin A1c, HF: heart failure, HFrEF: heart failure with reduced ejection fraction, HDL: high density lipoprotein, HTN: hypertension, INR: international normalized ratio, LDL: low density lipoprotein, TG: triglycerides, Tchol: total cholesterol, UTI: urinary tract infection, VTE: venous thromboembolism. | | | | | |

### Appendix 3 Outcomes in common conditions/Minor ailments (14 out of 43)

| **Disease area** | **Setting** | **Type of prescribing** | **Taxonomy core area** | **Domain** | **References** |
| --- | --- | --- | --- | --- | --- |
| **Clinical outcomes** | | | | | |
| 1. Clinical cure/ symptom resolution or improvement (9 out of 14) | | | | | |
| Minor ailments (no specified conditions) | Primary care | Independent | Physiological or clinical | 2–24: Physiological/clinical | Paudyal et al. (2013); Piraux et al. (2024); Faruquee CF (2015); Leong (2021) |
| Minor ailments (no specified conditions) | Primary care | Not reported/Unclear | Physiological or clinical | 2–24: Physiological/clinical | Curley et al. (2016) |
| Minor ailments (Musculoskeletal pain, cough and colds, eye irritation, allergic rhinitis, dyspepsia, constipation, diarrhoea, ear problems) | Primary care | Independent | Physiological or clinical | 2–24: Physiological/clinical | Yusuff et al. (2021) |
| Minor ailments (head lice, headache, high temperature, constipation, diarrhoea, dyspepsia/ heartburn, ringworm, oral thrush, haemorrhoids, cough, earache, hay fever, nasal symptoms, seasonal allergies, sore throat, allergic rhinitis, vaginal thrush, URTI, acne, athlete’s foot, canker sore, cold sore, diaper rash, dysmenorrhea, sprain, impetigo, insect bites, eczema, folliculitis, Herpes simplex) | Primary and secondary care | Unclear | Physiological or clinical | 2–24: Physiological/clinical | Paudyal et al. (2018) |
| Uncomplicated urinary tract infection (UTI) or cystitis | Primary care | Independent | Physiological or clinical | 2–24: Physiological/clinical | Wu JH et al. (2021) |
| Uncomplicated urinary tract infection (UTI) or cystitis | Primary care | Patient Group Directions (PGDs) | Physiological or clinical | 2–24: Physiological/clinical | Wu JH et al. (2021); Badran et al. (2024) |
| Dyspepsia | Primary care | Unclear | Physiological or clinical | 2–24: Physiological/clinical | Curley et al. (2016) |
| Cold sores | Primary care | Independent | Physiological or clinical | 2–24: Physiological/clinical | Wu JH et al. (2021) |
| Head lice | Primary care | Unclear | Physiological or clinical | 2–24: Physiological/clinical | Curley et al. (2016) |
| 2. Treatment failure (2 out of 14) | | | | | |
| Cold sores | Primary care | Independent | Physiological or clinical | 2–24: Physiological/clinical | Wu JH et al. (2021) |
| Uncomplicated urinary tract infection (UTI) or cystitis | Primary care | Independent | Physiological or clinical | 2–24: Physiological/clinical | Wu JH et al. (2021) |
| Uncomplicated urinary tract infection (UTI) or cystitis | Primary care | Patient Group Directions (PGDs) | Physiological or clinical | 2–24: Physiological/clinical | Wu JH et al. (2021); Badran et al. (2024) |
| 3. Re-consultation with the pharmacist or referral to other health care providers (5 out of 14) | | | | | |
| Minor ailments (no specified conditions) | Primary care | Independent prescribing | Resource use | 36: Need for further intervention | Paudyal et al. (2013); Leong (2021) |
| Minor ailments (head lice, headache, high temperature, constipation, diarrhoea, dyspepsia/ heartburn, ringworm, oral thrush, haemorrhoids, cough, earache, hay fever, nasal symptoms, seasonal allergies, sore throat, allergic rhinitis, vaginal thrush, URTI, acne, athlete’s foot, canker sore, cold sore, diaper rash, dysmenorrhea, sprain, impetigo, insect bites, eczema, folliculitis, Herpes simplex) | Primary and secondary care | Unclear | Resource use | 36: Need for further intervention | Paudyal et al. (2018) |
| Chronic obstructive pulmonary disease (COPD) exacerbation | Primary care | Patient Group Directions (PGDs) | Resource use | 36: Need for further intervention | Badran et al. (2024) |
| Uncomplicated urinary tract infection (UTI) or cystitis | Primary care | Patient Group Directions (PGDs) | Resource use | 36: Need for further intervention | Badran et al. (2024) |
| Dyspepsia | Primary care | Unclear | Resource use | 36: Need for further intervention | Curley et al. (2016) |
| Impetigo | Primary care | Patient Group Directions (PGDs) | Resource use | 36: Need for further intervention | Badran et al. (2024) |
| 4. Re-consultation with other health care providers/ other healthcare utilisation (2 out of 14) | | | | | |
| Minor ailments (no specified conditions) | Primary care | Independent prescribing | Resource use | 36: Need for further intervention | Paudyal et al. (2013) |
| Acute pharyngitis or sore throat, acute otitis media, acute bacterial sinusitis, chronic bacterial sinusitis | Primary care | Independent/Patient Group Directions (PGDs) | Resource use | 36: Need for further intervention | Wu JH et al. (2021) |
| Chronic obstructive pulmonary disease (COPD) exacerbation | Primary care | Patient Group Directions (PGDs) | Resource use | 36: Need for further intervention | Wu JH et al. (2021); |
| Uncomplicated urinary tract infection (UTI) or cystitis | Primary care | Patient Group Directions (PGDs) | Resource use | 36: Need for further intervention | Wu JH et al. (2021); |
| Uncomplicated urinary tract infection (UTI) or cystitis | Primary care | Independent | Resource use | 36: Need for further intervention | Wu JH et al. (2021) |
| **Drug-related and prescribing outcomes** | | | | | |
| 5. Prescribing rate/changes (1 out of 14) | | | | | |
| Acute pharyngitis or sore throat, acute otitis media, acute bacterial sinusitis, chronic bacterial sinusitis | Primary care | Independent/Patient Group Directions (PGDs) | Life impact | 32: Delivery of care (Process, implementation, and service outcomes) | Wu JH et al. (2021) |
| Uncomplicated urinary tract infection (UTI) or cystitis | Primary care | Patient Group Directions (PGDs) | Life impact | 32: Delivery of care (Process, implementation, and service outcomes) | Wu JH et al. (2021) |
| Chronic obstructive pulmonary disease (COPD) exacerbation | Primary care | Patient Group Directions (PGDs) | Life impact | 32: Delivery of care (Process, implementation, and service outcomes) | Wu JH et al. (2021) |
| 6. Guideline concordance/appropriateness of medications (3 out of 14) | | | | | |
| Uncomplicated urinary tract infection (UTI) or cystitis | Primary care | Patient Group Directions (PGDs) | Life impact | 32: Delivery of care (Appropriateness of treatment) | Wu JH et al. (2021); Badran et al. (2024) |
| Headache/back pain | Primary care | Unclear | Life impact | 32: Delivery of care (Appropriateness of treatment) | Curley et al. (2016) |
| Eye infection | Primary care | Unclear | Life impact | 32: Delivery of care (Appropriateness of treatment) | Curley et al. (2016) |
| 7. Patient adherence to medication (2 out of 14) |  |  |  |  |  |
| Minor ailments (head lice, headache, high temperature, constipation, diarrhoea, dyspepsia/ heartburn, ringworm, oral thrush, haemorrhoids, cough, earache, hay fever, nasal symptoms, seasonal allergies, sore throat, allergic rhinitis, vaginal thrush, URTI, acne, athlete’s foot, canker sore, cold sore, diaper rash, dysmenorrhea, sprain, impetigo, insect bites, eczema, folliculitis, Herpes simplex) | Primary care | Unclear | Life impact | 32: Delivery of care (Adherence/compliance) | Paudyal et al. (2018) |
| Cold sores | Primary care | Independent | Life impact | 32: Delivery of care (Adherence/compliance) | Wu JH et al. (2021) |
| Uncomplicated urinary tract infection (UTI) or cystitis | Primary care | Independent | Life impact | 32: Delivery of care (Adherence/compliance) | Wu JH et al. (2021) |
| 8. Adverse effects (4 out of 14) | | | | | |
| Minor ailments (no specified conditions) | Primary care | Independent | Adverse events | 38: Adverse events/effects | Piraux et al. (2024) |
| Minor ailments (head lice, headache, high temperature, constipation, diarrhoea, dyspepsia/ heartburn, ringworm, oral thrush, haemorrhoids, cough, earache, hay fever, nasal symptoms, seasonal allergies, sore throat, allergic rhinitis, vaginal thrush, URTI, acne, athlete’s foot, canker sore, cold sore, diaper rash, dysmenorrhea, sprain, impetigo, insect bites, eczema, folliculitis, Herpes simplex) | Primary care | Unclear | Adverse events | 38: Adverse events/effects | Paudyal et al. (2018) |
| Cold sores | Primary care | Independent | Adverse events | 38: Adverse events/effects | Wu JH et al. (2021) |
| Uncomplicated urinary tract infection (UTI) or cystitis | Primary care | Independent | Adverse events | 38: Adverse events/effects | Wu JH et al. (2021) |
| Uncomplicated urinary tract infection (UTI) or cystitis | Primary care | Patient Group Directions (PGDs) | Adverse events | 38: Adverse events/effects | Badran et al. (2024) |
| **Patient reported and experience outcomes** | | | | | |
| 9. General satisfaction (e.g. perceived ease and convenience) (6 out of 14) | | | | | |
| Minor ailments (no specified conditions) | Primary care | Independent | Life impact | 32: Delivery of care (Satisfaction/patient preference) | Jebara et al. (2018); Leong (2021) |
| Minor ailments (no specified conditions) | Primary care | Dependent | Life impact | 32: Delivery of care (Satisfaction/patient preference) | Walpola et al. (2024) |
| Acute pharyngitis or sore throat, acute otitis media, acute bacterial sinusitis, chronic bacterial sinusitis | Primary care | Independent/Patient Group Directions (PGDs) | Life impact | 32: Delivery of care (Satisfaction/patient preference) | Wu JH et al. (2021) |
| Group A streptococcal pharyngitis | Primary care | Collaborative | Life impact | 32: Delivery of care (Satisfaction/patient preference) | Walpola et al. (2024) |
| Uncomplicated urinary tract infection (UTI) or cystitis | Primary care | Independent | Life impact | 32: Delivery of care (Satisfaction/patient preference) | Wu JH et al. (2021) |
| Uncomplicated urinary tract infection (UTI) or cystitis | Primary care | Patient Group Directions (PGDs) | Life impact | 32: Delivery of care (Satisfaction/patient preference) | Wu JH et al. (2021); Badran et al. (2024) |
| Urinary tract infection (UTI) | Primary care | Dependent | Life impact | 32: Delivery of care (Satisfaction/patient preference) | Walpola et al. (2024) |
| Asthma | Primary care | Supplementary/independent | Life impact | 32: Delivery of care (Satisfaction/patient preference) | Jebara et al. (2018) |
| Chronic obstructive pulmonary disease (COPD) exacerbation | Primary care | Patient Group Directions (PGDs) | Life impact | 32: Delivery of care (Satisfaction/patient preference) | Wu JH et al. (2021); Badran et al. (2024) |
| Chronic obstructive pulmonary disease (COPD) | Primary care | Dependent | Life impact | 32: Delivery of care (Satisfaction/patient preference) | Walpola et al. (2024) |
| Impetigo | Primary care | Patient Group Directions (PGDs) | Life impact | 32: Delivery of care (Satisfaction/patient preference) | Badran et al. (2024) |
| Impetigo | Primary care | Dependent | Life impact | 32: Delivery of care (Satisfaction/patient preference) | Walpola et al. (2024) |
| Cold sores | Primary care | Independent | Life impact | 32: Delivery of care (Satisfaction/patient preference) | Wu JH et al. (2021) |
| Travel medicine | Primary care/ ambulatory | Collaborative/ Independent | Life impact | 32: Delivery of care (Satisfaction/patient preference) | Kc B et al. (2022) |
| 10. Quality of life/ general health status/ sleep and daily activity (1 out of 14) | | | | | |
| Minor ailments (head lice, headache, high temperature, constipation, diarrhoea, dyspepsia/ heartburn, ringworm, oral thrush, haemorrhoids, cough, earache, hay fever, nasal symptoms, seasonal allergies, sore throat, allergic rhinitis, vaginal thrush, URTI, acne, athlete’s foot, canker sore, cold sore, diaper rash, dysmenorrhea, sprain, impetigo, insect bites, eczema, folliculitis, Herpes simplex) | Primary and secondary care | Unclear | Life impact | 25-31: Functioning | Paudyal et al. (2018) |
| **Economic and other outcomes** | | | | | |
| 11. Cost of service delivery (9 out of 14) | | | | | |
| Minor ailments (no specified conditions) | Primary care | Independent | Resource use | 34: Economic | Paudyal et al. (2013); Piraux et al. (2024); Perraudin et al. (2016); Leong (2021) |
| Minor ailments (Musculoskeletal pain, cough and colds, eye irritation, allergic rhinitis, dyspepsia, constipation, diarrhoea, ear problems) | Primary care | Independent | Resource use | 34: Economic | Yusuff et al. (2021) |
| Acute pharyngitis or sore throat, acute otitis media, acute bacterial sinusitis, chronic bacterial sinusitis | Primary care | Independent/Patient Group Directions (PGDs) | Resource use | 34: Economic | Wu JH et al. (2021) |
| Uncomplicated urinary tract infection (UTI) or cystitis | Primary care | Patient Group Directions (PGDs) | Resource use | 34: Economic | Wu JH et al. (2021); Badran et al. (2024) |
| Chronic obstructive pulmonary disease (COPD) exacerbation | Primary care | Patient Group Directions (PGDs) | Resource use | 34: Economic | Wu JH et al. (2021); Badran et al. (2024) |
| Chronic obstructive pulmonary disease (COPD) exacerbation | Primary and secondary care | Non-medical switching | Resource use | 34: Economic | Krstic et al. (2021) |
| Peptic ulcer and gastroesophageal reflux diseases | Primary and secondary care | Non-medical switching | Resource use | 34: Economic | Krstic et al. (2021) |
| Cold sores | Primary care | Independent | Resource use | 34: Economic | Wu JH et al. (2021) |
| Impetigo | Primary care | Patient Group Directions (PGDs) | Resource use | 34: Economic | Badran et al. (2024) |
| Head lice | Primary care | Unclear | Resource use | 34: Economic | Curley et al. (2016) |
| 12. GP Workload (3 out of 14) | | | | | |
| Minor ailments (general) | Primary care | Independent | Resource use | 37: Societal/carer burden | Paudyal et al. (2013); Leong (2021) |
| Uncomplicated urinary tract infection (UTI) or cystitis | Primary care | Patient Group Directions (PGDs) | Resource use | 37: Societal/carer burden | Badran et al. (2024) |
| Chronic obstructive pulmonary disease (COPD) exacerbation | Primary care | Patient Group Directions (PGDs) | Resource use | 37: Societal/carer burden | Badran et al. (2024) |
| Impetigo | Primary care | Patient Group Directions (PGDs) | Resource use | 37: Societal/carer burden | Badran et al. (2024) |
| 13. Access to care (5 out of 14) | | | | | |
| Minor ailments (no specified conditions) | Primary care | Independent | Life impact | 32: Delivery of care (Acceptability and availability) | Piraux et al. (2024); Leong (2021) |
| Cold sores | Primary care | Independent | Life impact | 32: Delivery of care (Acceptability and availability) | Wu JH et al. (2021) |
| Acute pharyngitis or sore throat, acute otitis media, acute bacterial sinusitis, chronic bacterial sinusitis | Primary care | Independent/Patient Group Directions (PGDs) | Life impact | 32: Delivery of care (Acceptability and availability) | Wu JH et al. (2021) |
| Group A streptococcal pharyngitis | Primary care | Collaborative | Life impact | 32: Delivery of care (Acceptability and availability) | Walpola et al. (2024) |
| Uncomplicated urinary tract infection (UTI) or cystitis | Primary care | Independent | Life impact | 32: Delivery of care (Acceptability and availability) | Wu JH et al. (2021) |
| Uncomplicated urinary tract infection (UTI) or cystitis | Primary care | Patient Group Directions (PGDs) | Life impact | 32: Delivery of care (Acceptability and availability) | Wu JH et al. (2021); Badran et al. (2024) |
| Urinary tract infection (UTI) | Primary care | Dependent | Life impact | 32: Delivery of care (Acceptability and availability) | Walpola et al. (2024) |
| Chronic obstructive pulmonary disease (COPD) exacerbation | Primary care | Patient Group Directions (PGDs) | Life impact | 32: Delivery of care (Acceptability and availability) | Wu JH et al. (2021) |
| 14. Level of service activity (1 out of 14) | | | | | |
| Uncomplicated urinary tract infection (UTI) or cystitis | Primary care | Patient Group Directions (PGDs) | Resource use | 34: Economic | Wu JH et al. (2021) |

### Appendix 4 Outcomes in other acute and chronic conditions (35 out of 43)

| **Disease area** | **Setting** | **Type of**  **prescribing** | **Taxonomy core area** | **Domain** | **References** |
| --- | --- | --- | --- | --- | --- |
| **Clinical outcomes** | | | | | |
| 1. Mortality (5 out of 35) | | | | | |
| Ischaemic stroke or transient ischaemic attack | Primary care | Not reported/unclear | Death | 1: Mortality/survival | Weeks et al. (2016) |
| Cardiovascular (prior coronary artery disease) | Primary care | Not reported/unclear | Death | 1: Mortality/survival | Greer et al. (2018) |
| Cardiovascular (HF) | Primary care | Independent | Death | 1: Mortality/survival | Cao et al. (2021) |
| Cardiovascular (HF) | Primary care | Collaborative | Death | 1: Mortality/survival | Cao et al. (2021) |
| Cardiovascular (VTE/ Receiving anticoagulation) | Secondary care | Dependent | Death | 1: Mortality/survival | Poh et al. (2018) |
| Diabetes mellitus (poorly controlled) | Primary and secondary care | Not reported/unclear | Death | 1: Mortality/survival | Greer et al. (2018); Faruquee et al. (2015) |
| CKD (moderate to severe CKD (eGFR<45)) | Primary care | Not reported/unclear | Death | 1: Mortality/survival | Greer et al. (2018) |
| Geriatrics (older people with complex needs) | Primary care | Independent | Death | 1: Mortality/survival | Greer et al. (2018) |
| 2. Clinical effectiveness (e.g. BP and glycaemic Improvement/control, SBP/HbA1c/ LDL goals attainments, major CV events risk reduction, prevention of unplanned pregnancy, HIV prophylaxis) (15 out of 35) | | | | | |
| Ischaemic stroke or transient ischaemic attack | Primary care | Not reported/unclear | Physiological or clinical | 2–24: Physiological/clinical | Weeks et al. (2016) |
| Cardiovascular | Primary care | Not reported/unclear | Physiological or clinical | 2–24: Physiological/clinical | Weeks et al. (2016); Yuan et al. (2019); Faruquee et al. (2015) |
| Cardiovascular | Primary care | Independent | Physiological or clinical | 2–24: Physiological/clinical | Varas-Doval et al. (2021); Leong (2021);  Weeks et al. (2016) |
| Cardiovascular | Secondary care | Dependent | Physiological or clinical | 2–24: Physiological/clinical | Poh et al. (2018) |
| Cardiovascular | Secondary care | Collaborative | Physiological or clinical | 2–24: Physiological/clinical | Poh et al. (2018) |
| Cardiovascular | Secondary care | Supplementary | Physiological or clinical | 2–24: Physiological/clinical | Poh et al. (2018) |
| Diabetes mellitus | Primary care | Dependent | Physiological or clinical | 2–24: Physiological/clinical | Motlohi et al. (2023); Faruquee et al. (2015) |
| Diabetes mellitus | Primary care | Independent | Physiological or clinical | 2–24: Physiological/clinical | Motlohi et al. (2023); Leong (2021) |
| Diabetes mellitus | Primary and/ or secondary care | Collaborative | Physiological or clinical | 2–24: Physiological/clinical | Rotta et al. (2015); Wubben et al. (2012); Poh et al. (2018) |
| Diabetes mellitus | Secondary care | Supplementary | Physiological or clinical | 2–24: Physiological/clinical | Poh et al. (2018) |
| Diabetes mellitus | Primary and/ or secondary care | Not reported/unclear | Physiological or clinical | 2–24: Physiological/clinical | Weeks et al. (2016); Greer et al. (2018); Lum et al. (2023); Faruquee et al. (2015) |
| CKD | Primary care | Independent | Physiological or clinical | 2–24: Physiological/clinical | Ardavani et al. (2023) |
| CKD | Primary care | Collaborative | Physiological or clinical | 2–24: Physiological/clinical | Ardavani et al. (2023) |
| CKD | Primary care | Not reported/unclear | Physiological or clinical | 2–24: Physiological/clinical | Greer et al. (2018) |
| Mental health | Primary and secondary care | Dependent | Physiological or clinical | 2–24: Physiological/clinical | Rotta et al. (2015); Finley et al. (2023) |
| Mental health (Depression) | Primary care | Not reported/unclear | Physiological or clinical | 2–24: Physiological/clinical | Greer et al. (2018) |
| Chronic pain | Primary and/ or secondary care | Independent and Supplementary | Physiological or clinical | 2–24: Physiological/clinical | Shrestha et al. (2021); Weeks et al. (2016) |
| Gyn (hormonal contraception service) | Primary care | Patient Group Direction/Collaborative prescribing agreement | Physiological or clinical | 2–24: Physiological/clinical | Buckingham et al. (2021) |
| 3. Healthcare utilisation outcomes (e.g. all-cause hospitalisation, HF hospitalisation, Urgent Care/Emergency Room Visit) ((5 out of 35) | | | | | |
| Cardiovascular | Primary care | Not reported/unclear | Resource use | 36: Need for further intervention | Weeks et al. (2016) |
| Diabetes mellitus | Primary and/ or secondary care | Not reported/unclear | Resource use | 36: Need for further intervention | Lum et al. (2023) |
| Cardiovascular | Primary care | Independent/ Collaborative | Resource use | 36: Need for further intervention | Cao et al. (2021) |
| Mental health | Secondary care | Collaborative | Resource use | 36: Need for further intervention | Alshammari et al. (2023) |
| Geriatrics | Primary care | Independent | Resource use | 36: Need for further intervention | Liu et al. (2024) |
| **Drug-related and prescribing outcomes** | | | | | |
| 4. Prescribing pattern (rates and changes) (6 out of 35) | | | | | |
| Cardiovascular | Primary care | Not reported/unclear | Life impact | 32: Delivery of care (Process, implementation, and service outcomes) | Weeks et al. (2016) |
| Cardiovascular | Primary care | Supplementary | Life impact | 32: Delivery of care (Process, implementation, and service outcomes) | Bhanbhro et al. (2011) |
| Cardiovascular | Primary care | Collaborative | Life impact | 32: Delivery of care (Process, implementation, and service outcomes) | Walpola et al. (2024) |
| Mental health | Primary | Dependent | Life impact | 32: Delivery of care (Process, implementation, and service outcomes) | Rotta et al. (2015); Finley et al. (2023) |
| Mental health | Primary care | Collaborative | Life impact | 32: Delivery of care (Process, implementation, and service outcomes) | Walpola et al. (2024) |
| Diabetes mellitus | Primary and secondary care | Not reported/unclear | Life impact | 32: Delivery of care (Process, implementation, and service outcomes) | Lum et al. (2023) |
| Substance use disorder | Primary care | Collaborative/Independent | Life impact | 32: Delivery of care (Process, implementation, and service outcomes) | Walpola et al. (2024) |
| Gyn (hormonal contraception service) | Primary care | Dependent/Independent | Life impact | 32: Delivery of care (Process, implementation, and service outcomes) | Walpola et al. (2024) |
| 5. Guideline concordance/ appropriateness of medications (12 out of 35) | | | | | |
| Cardiovascular | Primary care | Not reported/unclear | Life impact | 32: Delivery of care (Appropriateness of treatment) | Greer et al. (2018) |
| Cardiovascular | Primary care | Independent | Life impact | 32: Delivery of care (Appropriateness of treatment) | Cao et al. (2021) |
| Cardiovascular | Primary care | Collaborative | Life impact | 32: Delivery of care (Appropriateness of treatment) | Cao et al. (2021) |
| Cardiovascular | Secondary care | Dependent | Life impact | 32: Delivery of care (Appropriateness of treatment) | Poh et al. (2018) |
| Cardiovascular | Secondary care | Supplementary | Life impact | 32: Delivery of care (Appropriateness of treatment) | Poh et al. (2018) |
| Geriatrics | Primary care | Dependent | Life impact | 32: Delivery of care (Appropriateness of treatment) | Rotta et al. (2015) |
| Geriatrics | Primary care | Independent | Life impact | 32: Delivery of care (Appropriateness of treatment) | Liu et al. (2024) |
| Oncology | Primary and secondary care | Collaborative | Life impact | 32: Delivery of care (Appropriateness of treatment) | Passey et al. (2023) |
| Opioid Analgesic Dependence | Primary care | Independent | Life impact | 32: Delivery of care (Appropriateness of treatment) | Jordan et al. (2021) |
| Perioperative settings | Secondary care | Independent | Life impact | 32: Delivery of care (Appropriateness of treatment) | Naseralallah et al. (2024); Noblet et al. (2018) |
| Perioperative settings | Secondary care | Supplementary | Life impact | 32: Delivery of care (Appropriateness of treatment) | Noblet et al. (2018); Poh et al. (2018) |
| Perioperative settings | Secondary care | Not reported/unclear | Life impact | 32: Delivery of care (Appropriateness of treatment) | Weeks et al. (2016) |
| ED patients | Secondary care | Independent | Life impact | 32: Delivery of care (Appropriateness of treatment) | Atey et al. (2023) |
| ED patients | Secondary care | Collaborative/ Supplementary | Life impact | 32: Delivery of care (Appropriateness of treatment) | Atey et al. (2023) |
| Mental health | Primary and secondary care | Dependent | Life impact | 32: Delivery of care (Appropriateness of treatment) | Finley et al. (2023) |
| 6. Adverse effects (8 out of 35) | | | | | |
| Ischaemic stroke or transient ischaemic attack | Primary care | Not reported/unclear | Adverse events | 38: Adverse events/effects | Weeks et al. (2016) |
| Cardiovascular | Primary care | Not reported/unclear | Adverse events | 38: Adverse events/effects | Weeks et al. (2016); Greer et al. (2018) |
| Cardiovascular | Primary care | Independent | Adverse events | 38: Adverse events/effects | Cao et al. (2021); Weeks et al. (2016) |
| Cardiovascular | Primary care | Collaborative | Adverse events | 38: Adverse events/effects | Cao et al. (2021) |
| Cardiovascular | Secondary care | Dependent | Adverse events | 38: Adverse events/effects | Poh et al. (2018) |
| Diabetes mellitus | Primary and secondary care | Not reported/unclear | Adverse events | 38: Adverse events/effects | Greer et al. (2018); Lum et al. (2023) |
| CKD | Primary care | Not reported/unclear | Adverse events | 38: Adverse events/effects | Greer et al. (2018) |
| Oncology | Primary and secondary care | Collaborative | Adverse events | 38: Adverse events/effects | Passey et al. (2023) |
| Chronic pain | Primary and secondary care | Not reported | Adverse events | 38: Adverse events/effects | Shrestha et al. (2021) |
| Geriatrics | Primary care | Independent | Adverse events | 38: Adverse events/effects | Liu et al. (2024) |
| 7. Patient adherence to medication (7 out of 35) | | | | | |
| Ischaemic stroke or transient ischaemic attack | Primary care | Not reported/unclear | Life impact | 32: Delivery of care (Adherence/compliance) | Weeks et al. (2016); Greer et al. (2018) |
| Cardiovascular | Primary care | Not reported/unclear | Life impact | 32: Delivery of care (Adherence/compliance) | Weeks et al. (2016) |
| Cardiovascular | Primary care | Independent | Life impact | 32: Delivery of care (Adherence/compliance) | Ali et al. (2024); Weeks et al. (2016) |
| Diabetes mellitus | Primary care | Dependent | Life impact | 32: Delivery of care (Adherence/compliance) | Ali et al. (2024) |
| Diabetes mellitus | Primary and/or secondary care | Not reported/unclear | Life impact | 32: Delivery of care (Adherence/compliance) | Weeks et al. (2016); Greer et al. (2018); Lum et al. (2023) |
| Mental health | Primary and secondary care | Dependent | Life impact | 32: Delivery of care (Adherence/compliance) | Rotta et al. (2015); Finley et al. (2023) |
| Mental health (depression) | Primary care | Not reported/unclear | Life impact | 32: Delivery of care (Adherence/compliance) | Weeks et al. (2016); Greer et al. (2018) |
| Oncology | Primary and secondary care | Collaborative | Life impact | 32: Delivery of care (Adherence/compliance) | Passey et al. (2023) |
| **Patient reported and experience outcomes** | | | | | |
| 8. General satisfaction (e.g. perceived ease and convenience) (18 out of 35) | | | | | |
| Ischaemic stroke or transient ischaemic attack | Primary care | Not reported/unclear | Life impact | 32: Delivery of care (Satisfaction/patient preference) | Weeks et al. (2016); Greer et al. (2018) |
| Cardiovascular | Primary care | Not reported/unclear | Life impact | 32: Delivery of care (Satisfaction/patient preference) | Weeks et al. (2016) |
| Cardiovascular | Primary and/ or secondary care | Dependent | Life impact | 32: Delivery of care (Satisfaction/patient preference) | Famiyeh et al. (2016); Poh et al. (2018) |
| Cardiovascular | Primary and/ or secondary care | Independent | Life impact | 32: Delivery of care (Satisfaction/patient preference) | Famiyeh et al. (2016); Walpola, et al. (2024) |
| Cardiovascular | Primary and/ or secondary care | Supplementary | Life impact | 32: Delivery of care (Satisfaction/patient preference) | Jebara et al. (2018); Bhanbhro et al. (2011) |
| Cardiovascular | Primary care | Collaborative | Life impact | 32: Delivery of care (Satisfaction/patient preference) | Walpola et al. (2024) |
| Respiratory | Primary and secondary care | Dependent | Life impact | 32: Delivery of care (Satisfaction/patient preference) | Famiyeh et al. (2016); |
| Respiratory | Primary and secondary care | Supplementary | Life impact | 32: Delivery of care (Satisfaction/patient preference) | Jebara et al. (2018) |
| Diabetes mellitus | Primary and secondary care | Dependent | Life impact | 32: Delivery of care (Satisfaction/patient preference) | Famiyeh et al. (2016); |
| Diabetes mellitus | Primary and / or secondary care | Independent | Life impact | 32: Delivery of care (Satisfaction/patient preference) | Famiyeh et al. (2016); Motlohi et al. (2023); |
| Diabetes mellitus | Primary and secondary care | Supplementary | Life impact | 32: Delivery of care (Satisfaction/patient preference) | Jebara et al. (2018) |
| Diabetes mellitus | Primary care | Collaborative | Life impact | 32: Delivery of care (Satisfaction/patient preference) | Rotta et al. (2015); Wubben et al. (2012) |
| Diabetes mellitus | Primary and secondary | Not reported/unclear | Life impact | 32: Delivery of care (Satisfaction/patient preference) | Greer et al. (2018); Lum et al. (2023) |
| CKD | Primary care | Collaborative | Life impact | 32: Delivery of care (Satisfaction/patient preference) | Ardavani et al. (2023) |
| Mental health | Primary and /or Secondary care | Dependent | Life impact | 32: Delivery of care (Satisfaction/patient preference) | Famiyeh et al. (2016); Rotta et al. (2015); Finley et al. (2023) |
| Mental health | Primary and /or Secondary care | Collaborative | Life impact | 32: Delivery of care (Satisfaction/patient preference) | Alshammari et al. (2023); Walpola, et al. (2024) |
| Mental health | Secondary care | Supplementary | Life impact | 32: Delivery of care (Satisfaction/patient preference) | Jebara et al. (2018) |
| Mental health (depression) | Primary care | Not reported/unclear | Life impact | 32: Delivery of care (Satisfaction/patient preference) | Greer et al. (2018) |
| Oncology and pain | Primary and secondary care | Dependent | Life impact | 32: Delivery of care (Satisfaction/patient preference) | Famiyeh et al. (2016); |
| Oncology and pain | secondary care | Supplementary | Life impact | 32: Delivery of care (Satisfaction/patient preference) | Jebara et al. (2018) |
| Oncology | Primary and secondary care | Collaborative | Life impact | 32: Delivery of care (Satisfaction/patient preference) | Passey et al. (2023) |
| Substance use disorder | Primary and/or secondary care | Dependent and Independent | Life impact | 32: Delivery of care (Satisfaction/patient preference) | Famiyeh et al. (2016); Jebara et al. (2018); Walpola et al. (2024) |
| General practice | Primary and secondary care | Dependent | Life impact | 32: Delivery of care (Satisfaction/patient preference) | Famiyeh et al. (2016); |
| General practice | Primary and secondary care | Independent | Life impact | 32: Delivery of care (Satisfaction/patient preference) | Famiyeh et al. (2016); |
| Gyn (hormonal contraception | Primary care | Dependent/Collaborative and independent | Life impact | 32: Delivery of care (Satisfaction/patient preference) | Kennedy et al. (2019); Walpola et al. (2024) |
| HIV preexposure prophylaxis | Primary care | Collaborative | Life impact | 32: Delivery of care (Satisfaction/patient preference) | Walpola et al. (2024); Zhao et al. (2021) |
| Chronic pain | Primary and/ or secondary care | Independent and Supplementary | Life impact | 32: Delivery of care (Satisfaction/patient preference) | Shrestha et al. (2021); Weeks et al. (2016) |
| Geriatrics | Primary care | Not reported/unclear | Life impact | 32: Delivery of care (Satisfaction/patient preference) | Greer et al. (2018) |
| 9. Quality of life/ general health status/ Disability (7 out of 35) | | | | | |
| Cardiovascular | Primary care | Not reported/unclear | Life impact | 25-31: Functioning | Weeks et al. (2016); Greer et al. (2018) |
| Diabetes mellitus | Primary and secondary care | Not reported/unclear | Life impact | 25-31: Functioning | Weeks et al. (2016); Greer et al. (2018); Lum et al. (2023); Faruquee et al. (2015) |
| Chronic pain | Primary and/ or secondary care | Independent and Supplementary | Life impact | 25-31: Functioning | Noblet et al. (2018); Shrestha et al. (2021); Weeks et al. (2016) |
| CKD | Primary care | Not reported/unclear | Life impact | 25-31: Functioning | Greer et al. (2018) |
| Mental health (depression) | Primary care | Not reported/unclear | Life impact | 25-31: Functioning | Greer et al. (2018) |
| Geriatrics | Primary care | Not reported/unclear | Life impact | 25-31: Functioning | Greer et al. (2018) |
| Geriatrics | Primary care | Independent | Life impact | 25-31: Functioning | Liu et al. (2024) |
| **Resource use and economic outcomes** | | | | | |
| 10. Access to care (9 out of 35) | | | | | |
| Cardiovascular | Primary care | Not reported/unclear | Life impact | 32: Delivery of care (Acceptability and availability) | Greer et al. (2018) |
| Cardiovascular | Primary and secondary care | Dependent | Life impact | 32: Delivery of care (Acceptability and availability) | Famiyeh et al. (2016); |
| Cardiovascular | Primary and secondary care | Independent | Life impact | 32: Delivery of care (Acceptability and availability) | Famiyeh et al. (2016); |
| Cardiovascular | Primary and/ or secondary care | Supplementary | Life impact | 32: Delivery of care (Acceptability and availability) | Jebara et al. (2018); Bhanbhro et al. (2011) |
| Cardiovascular | Primary care | Collaborative | Life impact | 32: Delivery of care (Acceptability and availability) | Walpola, et al. (2024) |
| Respiratory | Primary and secondary care | Dependent | Life impact | 32: Delivery of care (Acceptability and availability) | Famiyeh et al. (2016); |
| Respiratory | Primary and secondary care | Supplementary | Life impact | 32: Delivery of care (Acceptability and availability) | Jebara et al. (2018) |
| Diabetes mellitus | Primary and secondary care | Dependent | Life impact | 32: Delivery of care (Acceptability and availability) | Famiyeh et al. (2016); |
| Diabetes mellitus | Primary and secondary care | Independent | Life impact | 32: Delivery of care (Acceptability and availability) | Famiyeh et al. (2016); |
| Diabetes mellitus | Primary and secondary care | Supplementary | Life impact | 32: Delivery of care (Acceptability and availability) | Jebara et al. (2018) |
| Diabetes mellitus | Primary and secondary care | Not reported/unclear | Life impact | 32: Delivery of care (Acceptability and availability) | Greer et al. (2018) |
| Mental health | Secondary care | Dependent | Life impact | 32: Delivery of care (Acceptability and availability) | Famiyeh et al. (2016); |
| Mental health | Secondary care | Supplementary | Life impact | 32: Delivery of care (Acceptability and availability) | Jebara et al. (2018) |
| Mental health | Primary care | Collaborative | Life impact | 32: Delivery of care (Acceptability and availability) | Walpola, et al. (2024) |
| Mental health (depression) | Primary care | Not reported/unclear | Life impact | 32: Delivery of care (Acceptability and availability) | Greer et al. (2018) |
| Oncology and pain | Primary and secondary care | Dependent | Life impact | 32: Delivery of care (Acceptability and availability) | Famiyeh et al. (2016); |
| Oncology and pain | secondary care | Supplementary | Life impact | 32: Delivery of care (Acceptability and availability) | Jebara et al. (2018) |
| Oncology | Primary and secondary care | Collaborative | Life impact | 32: Delivery of care (Acceptability and availability) | Passey et al. (2023) |
| Substance use disorder | Primary and/ or secondary care | Dependent and Independent/ Collaborative | Life impact | 32: Delivery of care (Acceptability and availability) | Famiyeh et al. (2016); Jebara et al. (2018); Walpola et al. (2024) |
| General practice | Primary and secondary care | Dependent | Life impact | 32: Delivery of care (Acceptability and availability) | Famiyeh et al. (2016); |
| General practice | Primary and secondary care | Independent | Life impact | 32: Delivery of care (Acceptability and availability) | Famiyeh et al. (2016); |
| HIV preexposure prophylaxis | Primary care | Collaborative | Life impact | 32: Delivery of care (Acceptability and availability) | Walpola et al. (2024); Zhao et al. (2021) |
| Geriatrics | Primary care | Independent | Life impact | 32: Delivery of care (Acceptability and availability) | Walpola et al. (2024) |
| Gyn (hormonal contraception service) | Primary care | Patient Group Direction/Collaborative and Independent | Life impact | 32: Delivery of care (Acceptability and availability) | Buckingham et al. (2021); Newlon et al. (2021); Walpola et al. (2024) |
| 11. Cost of service delivery (15 out of 35) | | | | | |
| Cardiovascular | Primary care | Not reported/unclear | Resource use | 34: Economic | Weeks et al. (2016); Greer et al. (2018) |
| Cardiovascular | Primary and/ or secondary care | Independent | Resource use | 34: Economic | Babashahi et al. (2023); Leong (2021) |
| Cardiovascular | Primary and secondary care | Non-medical switching | Resource use | 34: Economic | Krstic et al. (2021) |
| Diabetes mellitus | Primary and secondary care | Non-medical switching | Resource use | 34: Economic | Krstic et al. (2021) |
| Diabetes mellitus | Primary and secondary care | Independent | Resource use | 34: Economic | Leong (2021) |
| Diabetes mellitus | Primary and secondary | Not reported/unclear | Resource use | 34: Economic | Greer et al. (2018); Lum et al. (2023); Faruquee et al. (2015) |
| CKD | Primary care | Not reported/unclear | Resource use | 34: Economic | Greer et al. (2018) |
| Gyn (hormonal contraception service) | Primary care | Patient Group Direction/Collaborative Prescribing agreement and Independent | Resource use | 34: Economic | Buckingham et al. (2021); Newlon et al. (2021) |
| Mental health | Primary and secondary care | Dependent | Resource use | 34: Economic | Rotta et al. (2015); Finley et al. (2023) |
| Mental health | Primary and secondary care | Non-medical switching | Resource use | 34: Economic | Krstic et al. (2021) |
| Mental health (depression) | Primary care | Not reported/unclear | Resource use | 34: Economic | Greer et al. (2018) |
| Oncology | Primary and secondary care | Collaborative | Resource use | 34: Economic | Passey et al. (2023) |
| Chronic pain | Primary and/or care | Independent/ Supplementary | Resource use | 34: Economic | Perraudin et al. (2016); Dawoud et al. (2019); Babashahi et al. (2023); Leong (2021);Noblet et al. (2018) |
| Geriatrics | Primary care | Not reported/unclear | Resource use | 34: Economic | Greer et al. (2018) |
| 12. Resource use due to health care utilisation (2 out of 35) | | | | | |
| Cardiovascular | Primary care | Not reported/unclear | Resource use | 34-37: Resource use | Weeks et al. (2016) |
| Diabetes mellitus | Primary and/ or secondary care | Not reported/unclear | Resource use | 34-37: Resource use | Weeks et al. (2016); Lum et al. (2023) |
| Depression | Primary care | Not reported/unclear | Resource use | 34-37: Resource use | Weeks et al. (2016) |

### Appendix 5 Overview of results from the rapid review

A. **Outcomes for prescribing for minor ailments**

**Clinical outcomes**: symptom improvement or clinical cure (i.e. symptom resolution) was cited as an outcome of pharmacist prescribing in nine reviews [6, 10, 13, 21, 29, 30, 32, 39, 42]. For example, Wu et al. reported that community pharmacist prescribing of systemic antimicrobials for conditions such as uncomplicated urinary tract infections, acute pharyngitis, and chronic obstructive pulmonary disease exacerbations was effective in achieving clinical cure and complete symptom improvement [39]. Treatment failure, when the prescribed medication does not significantly or completely improve the symptoms of the index condition, was reported in two reviews [6, 39]. Six reviews assessing the impact of pharmacist prescribing in several minor aliments reported on referral back to GP (i.e. reviews by Curley et al., Paudyal et al. (2013), Badran et al, and Leong et al) [6, 10, 21, 30], re-consultation with the pharmacist (i.e. reviews by Curley et al. and Paudyal et al. (2018)) [10, 29] and/or re-consultation with other health care providers (e.g. GP) (Paudyal et al. (2013) and Wu et al.) [30, 39].

**Prescribing related outcomes**: Four reviews reported on adverse effects of the prescribed medications [6, 29, 32, 39], and two reviews on patient adherence to medications [29, 39]. Three reviews assessed pharmacists’ adherence to clinical management/prescribing guidelines or protocols, and the quality of prescribing (e.g. appropriate drug selection at appropriate dose and frequency, addressing omitted medications, and ensuring medicines are properly discontinued) [6, 10, 39]. Wu and colleagues conducted a systematic review on community pharmacist prescribing of antimicrobials for uncomplicated urinary tract infections, acute pharyngitis, and chronic obstructive pulmonary disease exacerbations, examining it from the perspective of antimicrobial stewardship, and reported on antimicrobial prescribing rate or utilisation [39].

**Patient reported and experience outcomes**: Patient satisfaction and experience were assessed in six reviews exploring the impact of pharmacist prescribing [6, 16, 18, 21, 37, 39]. For instance, the systematic review by Jebara and colleagues examined stakeholders' views and experiences of pharmacist prescribing for minor ailments in primary care settings [16]. The study explored patient experience and satisfaction through questionnaires addressing various aspects, including access to medicines, quality of care, knowledge and adherence to prescribed medicines, the patient–professional relationship, and patients' perceptions of condition control under pharmacist-led services. Five reviews also highlighted the role of pharmacist prescribers in improving access to medicines and expediting care [6, 21, 32, 37, 39]. Walpola and colleagues reported that pharmacist prescribers were able to implement medication changes and issue prescriptions for minor ailments and other conditions more quickly than physicians [37]. Common reasons for choosing a pharmacist prescriber included the ease of accessing a pharmacist, avoiding delays associated with physician appointments, and the convenience of receiving care when medical clinics were unavailable or when patients lacked a regular physician. Additionally, Paudyal et al. (2018) reviewed the impact of pharmacist prescribing for minor aliments on health-related quality of life, general health status and the impact of illness on sleep and daily activity [29].

**Economic and related outcomes**: Economic outcomes such as cost of service delivery were among the most commonly cited outcomes used to assess impact of pharmacist prescribing for minor ailments in the retrieved reviews (n = 9/14) [6, 10, 20, 21, 29-32, 39]. For instance, Perraudin et al. found that professional pharmacy services to enhance access to minor ailment scheme medicines had evidence supporting their cost-effectiveness and cost minimisation in the UK [31]. Wu and colleagues highlighted the cost-effectiveness of pharmacist prescribing, with significant cumulative cost savings and a positive return on investment five years post-implementation in a broader provincial ambulatory ailments program [39]. Wu et al. also evaluated the level of service activity by measuring the number of service claims [39]. Moreover, three reviews assessed the impact on GP workload due to task shifting of common conditions consultations [6, 21, 30].

B. **Outcomes for prescribing in other contexts**

Outcomes evaluating the impact of pharmacist prescribing beyond minor ailments were also categorised into four groups: clinical outcomes, drug-related and prescribing outcomes, patient-reported and experience outcomes, and economic and other related outcomes. A new clinical outcome, mortality, was cited in five reviews as an outcome evaluating the impact of independent pharmacist prescribing, which was not addressed in the context of common conditions [9, 13, 15, 33, 38]. Notably, clinical effectiveness in independent prescribing corresponds to clinical cure and symptom improvement in common conditions, while healthcare utilisation could correspond with re-consultation or referral to pharmacists or other healthcare providers in reviews of common conditions. Healthcare utilisation highlights the use or presentations for further medical care—such as primary carer visits, all cause emergency department (ED) visits, condition ‐related ED visits or hospitalisations—as indicators of/ Proxy for clinical or safety outcomes (i.e. improving or worsening of the condition following pharmacist prescribing). For example, the review by Cao et al. showed a significant decrease in all-cause hospitalisation following pharmacist-led medication titration- with independent authority or under a collaborative practice agreement- in ambulatory patients with Heart Failure [9].

Additionally, independent pharmacist prescribing demonstrates a broader scope in drug-related and prescribing outcomes, particularly with the inclusion of detailed prescribing patterns, which encompass prescribing rates and medication changes. Walpola and colleagues included three studies reporting an increase in the percentage of eligible patients receiving relevant medicines following introduction of pharmacist prescribing services for depression (+9 %), opioid use disorder (+3 %), and atherosclerotic cardiovascular disease (+4 %) [37]. Two studies reported an increase in total dispensing for emergency contraception (+102 %) and naloxone (+53 %) following implementation of pharmacist prescribing. In another systematic review by Finely et al., pharmacist dependent prescribing models in mental health improved prescribing patterns, most commonly reducing the dosage and absolute number of psychotropic drugs [14]. Moreover, Weeks et al. has synthesised evidence on non‐medical prescribing (including pharmacist) versus medical prescribing for acute and chronic disease management in primary and secondary care [38]. Pharmacist prescribing resulted in a higher initiation of new cardiovascular medicines (antihypertensive agents), more frequent dose changes, an increased number of medicines being discontinued, and a greater prescription rate of low-dose aspirin and statins compared to the usual care group.

Similar to minor ailments, patients’s experience and satisfaction was highly cited as an outcome evaluating the impact of independent pharmacist prescribing across primary and secondary care. Famiyeh and colleagues conducted a scoping review exploring patient experiences with pharmacist prescribing in primary and secondary care settings, specifically within independent and dependent prescribing models [12]. The review extensively evaluated general satisfaction, perceived ease, and convenience associated with pharmacist-led care. It highlighted positive experiences among patients with chronic conditions (e.g. cardiovascular diseases, diabetes mellitus, respiratory diseases, substance misuse, oncology and pain related conditions, emphasising improvements and general satisfaction across key dimensions such as access to care, interpersonal communication, continuity and coordination, comprehensiveness of services, trust in healthcare providers, and patient-reported impacts of care.

Economic and other outcomes also show additional considerations, particularly resource use due to healthcare utilisation. This emphasised the measurement of health facility resource use driven by events such as hospitalisations, ED visits, and outpatient visit, as a proxy for economic evaluation. For example, Lum et al. found pharmacist‐led diabetes mellitus management resulted in improved medical resource consumption (outpatient visits, hospitalisations, ED visits) [23].

### Appendix 6 Included reviews
